# The potential clinical impact and cost-effectiveness of the updated COVID-19 mRNA Fall 2023 vaccines in the United States

**DOI:** 10.1101/2023.09.05.23295085

**Authors:** M Kohli, M Maschio, K Joshi, A Lee, K Fust, E Beck, N Van de Velde, MC Weinstein

## Abstract

**Objectives:** To assess the potential clinical impact and cost-effectiveness of COVID-19 mRNA vaccines updated for Fall 2023 in adults ≥18 years over a 1-year analytic time horizon (September 2023-August 2024).

**Methods:** A compartmental Susceptible-Exposed-Infected-Recovered model was updated to reflect COVID-19 in summer 2023. Numbers of symptomatic infections, COVID-19 related hospitalizations and deaths, and costs and quality-adjusted life-years (QALYs) gained were calculated using a decision tree model. The incremental cost-effectiveness ratio (ICER) of a Moderna updated mRNA Fall 2023 vaccine (Moderna Fall Campaign) was compared to no additional vaccination. Potential differences between the Moderna and the Pfizer-BioNTech Fall 2023 vaccines were examined.

**Results:** Base case results suggest the Moderna Fall Campaign would decrease the expected 64.2 million symptomatic infections by 7.2 million (11%) to 57.0 million. COVID-19-related hospitalizations and deaths are expected to decline by 343,000 (–29%) and 50,500 (–33%), respectively. The Moderna Fall Campaign would increase QALYs by 740,880 and healthcare costs by $5.7 billion relative to No Vaccine, yielding an ICER of $7,700 per QALY gained. Using a societal cost perspective, the ICER is $2,100. Sensitivity analyses suggest that vaccine effectiveness, COVID-19 incidence, hospitalization rates and costs drive cost-effectiveness. With a relative vaccine effectiveness (rVE) of Moderna versus Pfizer-BioNTech of 5.1% for infection and 9.8% for hospitalization, use of the Moderna vaccine is expected to prevent 24,000 more hospitalizations and 3,300 more deaths than the Pfizer-BioNTech vaccine.

**Limitations and Conclusions:** As COVID-19 becomes endemic, future incidence, including patterns of infection, are highly uncertain. Vaccine effectiveness of Fall 2023 vaccines is unknown, and it is unclear when a new variant that evades natural or vaccine immunity will emerge. Despite these limitations, the Moderna Fall 2023 vaccine can be considered cost-effective relative to no vaccine.

## Introduction

In May 2023, the World Health Organization (WHO) ended the COVID-19 global health emergency following 12 months of decreasing incidence of infections.^1^ The United States (US) federal government also ended its public health emergency on May 11, 2023.^2^ The World Health Organization (WHO) attributed the decrease in the risk of infection to increased levels of immunity achieved through both highly effective vaccines and previous infections. In the US, for example, Jones et al. report an increase in SARS-CoV-2 antibodies amongst blood donors at 68.4% from April – June 2021 to 96.4% from July – September 2022.^3^ Among those tested in the latter time period, 22.6% had antibodies from a previous infection alone, 26.1% had antibodies from vaccination alone, and 47.7% had hybrid immunity.

The use of COVID-19 vaccines has been evolving throughout the pandemic. In the US, the mRNA vaccines produced by Moderna and Pfizer-BioNTech are preferred, and a third protein sub-unit vaccine manufactured by Novavax is also available.^4^ As SARS-CoV-2 evolved from the ancestral strain (Wuhan-hu-1) into the Omicron variants that have circulated since January 2022,^5^ the COVID-19 vaccines have been updated. New bivalent COVID-19 mRNA vaccines containing antigens to both the ancestral strain and the Omicron BA.4/BA.5 sub-variants were developed. In September 2022, the Centers for Disease Control (CDC) recommended that all individuals aged 6 months and above receive one dose of these bivalent vaccines.^6^ By the end of January 2023, XBB sub-variants had began to dominate globally, and initial studies reported that the bivalent COVID-19 vaccines have low effectiveness against these sub-variants.^7,8^ Therefore, in May 2023, the WHO Technical Advisory Group on COVID-19 Vaccine Composition recommended that COVID-19 vaccines be updated once more to monovalent versions with an XBB subvariant. In the United States (US), in June 2023, the Vaccines and Related Biological Products Advisory Committee (VRBPAC) of the Food and Drug Administration (FDA) specified that COVID-19 vaccines in the US be updated to monovalent versions with the XBB.1.5 sub-lineage of Omicron.^9^ As of the end of July 2023, XBB sub-lineages accounted for 98% of COVID-19 infections in the US.^10^

During the pandemic phase of COVID-19, the few formal economic evaluations conducted of the vaccines concluded that they were a cost-effective intervention.^11^ Previously, we published an analysis of the potential clinical impact of the bivalent boosters in the Fall of 2022 in the US using a mathematical model of infection dynamics.^12^ We did not conduct an economic assessment, and the incidence of COVID-19 infections has since declined. While the CDC continued to distribute free COVID-19 vaccines in May 2023, the funding of COVID-19 vaccines in the US is transitioning from the pandemic procurement process to the standard processes for childhood and adult vaccines. Given these changes to the epidemiological, clinical and financial dimensions of COVID-19 vaccines, it is important to assess both the ongoing clinical and economic impact of additional COVID-19 doses. Specifically, it will be important to determine if use of COVID-19 vaccines is still cost-effective from both a societal and a health care system perspective.

The objective of this analysis was to assess the potential clinical impact and cost-effectiveness of the mRNA COVID-19 vaccines updated for Fall 2023 in persons ages 18 years and above, using a mathematical model. Outcomes were estimated both with and without a Moderna updated Fall 2023 COVID-19 vaccine campaign. In addition, we explore the potential differences between the mRNA COVID-19 vaccines manufactured by Moderna and Pfizer-BioNTech. The formulation of these two mRNA COVID-19 vaccines differ in multiple points, such as dosage and delivery system (lipid nanoparticles), and observational studies have demonstrated that these differences impact vaccine effectiveness. ^13-18^ Therefore, as an additional analysis, we compare the potential benefits of a Fall 2023 vaccination campaign with the Moderna updated COVID-19 mRNA vaccine to one with the Pfizer-BioNTech updated COVID-19 mRNA vaccine.

## Methods

### Overview

For this analysis, two sets of comparisons were conducted across the 1-year analytic time horizon of September 2023 to August 2024. First, vaccination of those 18 years and older with the Moderna updated COVID-19 mRNA Fall 2023 vaccine (Moderna Fall Campaign) was compared to no additional COVID-19 vaccination in Fall 2023 (No Vaccine Fall Campaign). Second, the Moderna Fall Campaign was compared to vaccination of those 18 years and older with the Pfizer-BioNTech updated COVID-19 mRNA Fall 2023 vaccine (Pfizer-BioNTech Fall Campaign).

A Susceptible-Exposed-Infected-Recovered (SEIR) model was used to estimate the total number of infections during the time horizon. A consequences decision tree was then used to calculate the numbers of symptomatic infections, COVID-19 related hospitalizations, COVID-19 related deaths, and associated costs and quality-adjusted life-years (QALYs) gained for each vaccination strategy. The incremental cost-effectiveness ratio (ICER) comparing the two vaccination strategies was calculated as the difference in costs between the Moderna Fall Campaign and the No Vaccine Fall Campaign divided by the difference in QALYs (incremental cost per QALY gained).

### SEIR Dynamic Transmission Model

A previously developed compartmental SEIR model was updated to reflect the clinical and epidemiological situation in the summer of 2023. The model has been previously described in detail;^12^ in the updated model with all of the SEIR compartments are stratified by vaccination status as shown in Figure 1. To accommodate changes since the summer of 2022, one more stratum was added to represent the Fall 2023 vaccine. Individuals in the Susceptible (S) state move to the Exposed (E) state based on the force (incidence) of infection which is affected by the susceptible individual’s vaccination stratum. The Exposed (E) state represents an infection that is both asymptomatic and nontransmissible, and which lasts for an average of 3 days.^19^ The Infected (I) state represents a contagious infection that may be clinically symptomatic or asymptomatic for an average of 7 days.^19,20^ Finally, individuals in the Recovered (R) states have natural immunity to infection, and the rate of movement back to the Susceptible (S) states represents the waning of natural immunity. Anyone who is in the S or R states may receive a vaccine. Once individuals have received their primary series, they move into the “Vaccinated” stratum. They progress through the additional booster 1 and booster 2 strata over time as additional doses are received.

**Figure 1.**
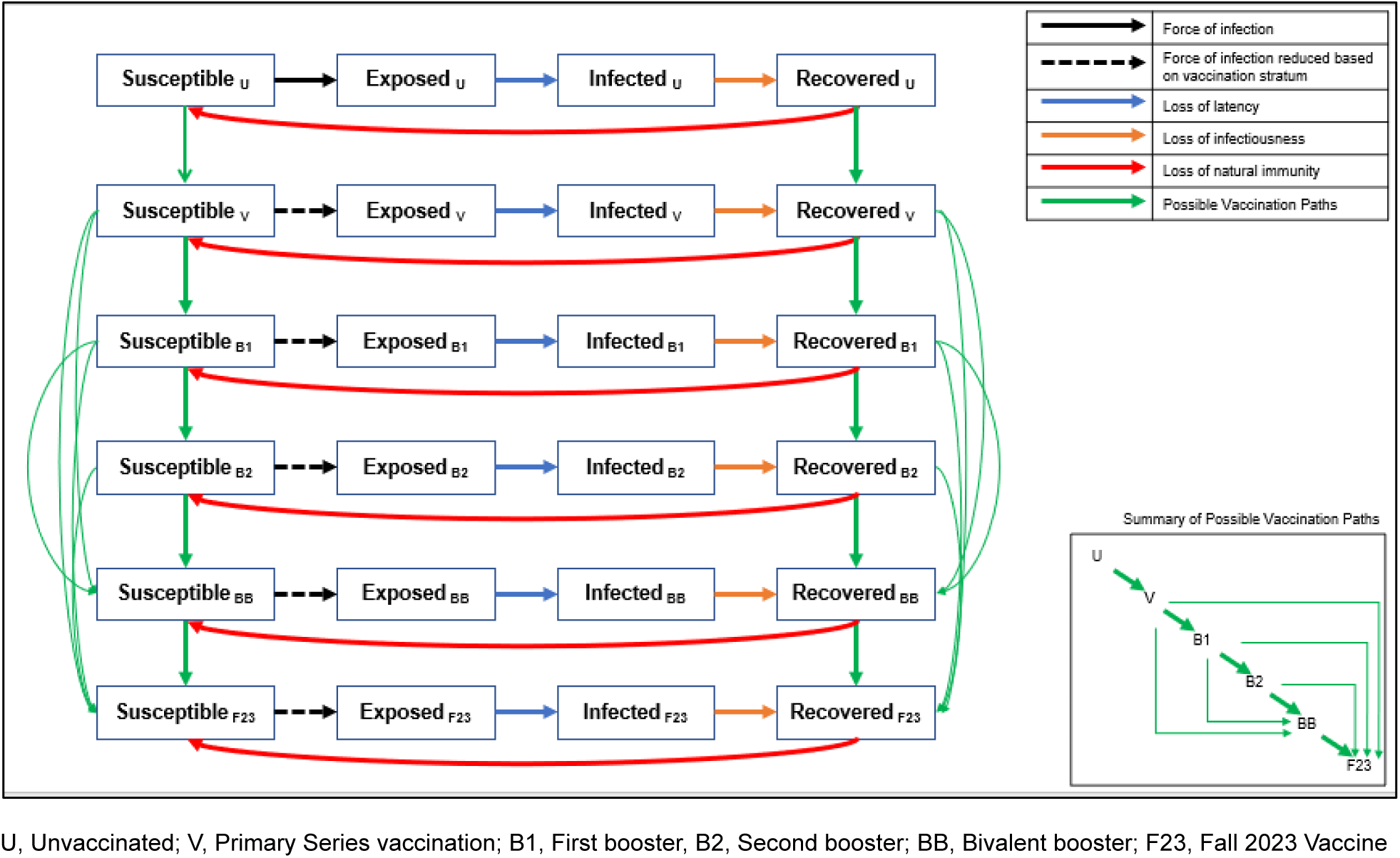
Model structure: compartments in the SEIR dynamic model of COVID-19 infection. Black arrows represent the movement between the susceptible and exposed compartments which is driven by the force of infection. The dashed black arrows indicate that the force of infection is modified by vaccination compared to the same transition in the unvaccinated stratum. Each vaccine stratum is associated with a unique vaccine effectiveness estimate. The blue arrows represent the loss of latency which means the infection becomes transmissible. The green arrows represent the loss of infectiousness which means the infection is cleared and natural immunity develops. The red arrows represent the loss of natural immunity following infection and transition back to the susceptible state. Loss of latency, loss of infectiousness and loss of natural immunity is the same for all vaccine strata but can change over time. The green arrows represent the possible vaccination points. The inset box is a stylized version that shows the possible vaccination points more clearly. Please see the Technical Appendix for the mathematical equations driving each transition.

Consistent with CDC policies, from September 2022 to August 2023, any person who had received at least their primary series could receive the bivalent mRNA vaccine irrespective of subsequent boosters. This analysis assumes that, after August 2023, individuals who had received at least their primary series are eligible to receive an updated COVID-19 mRNA Fall 2023 vaccine, whether or not they have received monovalent or bivalent boosters. Differential equations that define the transitions in the model, and the associated parameter inputs are provided in the Technical Appendix.

In the SEIR model, vaccination reduces the incidence of asymptomatic or symptomatic infection. Vaccination can also reduce the risk of hospitalization for someone with a symptomatic infection. Each day, newly vaccinated individuals are assigned the initial vaccine effectiveness (VE) that is appropriate to the vaccine used and the variant that is circulating. The VE of previously vaccinated persons is reduced according to the waning rate. The average VE for a given day is calculated as a weighted average of the VE for the newly vaccinated individuals and the VE of those vaccinated in past days, weighted by the number of people in each of these groups. The VE against hospitalization, i.e., the reduction in the risk of hospitalization given infection for vaccinated versus unvaccinated persons is calculated for use in the consequences decision tree (see Technical Appendix).

### SEIR Model Time Periods

The model simulation is run from the emergence of COVID-19 in January 2020 until August 2024, with a time step of 1 day. There are two time periods considered in the model simulation. The first is the “burn-in” (or “model calibration”) time period, which runs from model simulation until August 2023. The burn-in time period is used to estimate the residual protection from previously received vaccines and the proportion of individuals with natural immunity (in the R compartments) at the end of August 2023. The second is the analysis time period, which runs from September 2023 to August 2024, aligns with the 1-year time horizon for the cost-effectiveness analysis. It is used to project the incidence of COVID-19 and the potential impact of Fall 2023 vaccines.

### Burn-in Period

The model simulation considers three variant time periods during the burn-in period: 1) the pre-Omicron period (January 31, 2020 – November 30, 2021); 2) the Omicron BA.1 period (December 1, 2021 – August 14, 2022); and 3) the Omicron BA.4/BA.5 period (August 15, 2022 – February 28, 2023). During variant time period 1, all vaccines administered were considered well-matched to circulating COVID-19 variants, so VE depends only on the time since vaccination. However, at the start of variant time periods 2 and 3, it is assumed that a new variant with immune escape has emerged. Therefore, the VE of any previously administered vaccine immediately declines, and a proportion of people immediately lose natural immunity and are moved from the R to the S compartments. Any newly administered poorly-matched vaccines also have lower initial VE compared to a well-matched vaccine.

During the burn-in period, vaccine uptake patterns vary by age group. The residual vaccine effectiveness (VE) protection at the end of the burn-in period varies by vaccine stratum because the timing of vaccine receipt of each booster is based on historical recommendations and coverage patterns.

The proportions of people in the R states depend on how many have previously developed an infection, which depends on social contact patterns and virus transmissibility. During the burn-in period, social contact patterns that drive risk of transmission were altered greatly during the pandemic through reduced mobility and risk reduction practices such as mask wearing. In addition, the transmissibility of the virus changed as new variants emerged and was modified by COVID-19 vaccines. While contact patterns and VE were estimated based on data, the transmissibility of the virus was derived through a calibration process described in the Technical Appendix. The purpose of the calibration process was to ensure that the model predicted a valid number of infections, ensuring that a reasonable proportion of people ended up in the R compartment on August 31, 2023. The transmissibility parameter was then held constant during the analysis time horizon.

### Burn-in Period: Alternative Infection Scenarios

In addition to the base case scenario, three additional calibrations were conducted in order to allow variation in the proportion of individuals with natural immunity and the amount of residual vaccine effectiveness at the end of the burn-in period. For the first alternative burn-in scenario, the waning rate of natural protection from a COVID-19 infection during Omicron was doubled and then the model was recalibrated to the same level as in the base case. For the second, the waning rate was reduced by half. A final alternative burn-in scenario was created in which a drop in VE was added on January 15, 2023 to mimic the emergence of XBB variants and reduce the residual VE accordingly. These changes ultimately lead to different incidence projections during the analysis time horizon. Further details are provided in the Technical Appendix.

### Analysis Time Horizon

To conduct the analyses, the SEIR model simulation was run multiple times, varying the vaccination characteristics (initial VE; waning of VE; vaccine coverage) for the Fall 2023 Vaccine campaigns (i.e., Moderna Fall Campaign, Pfizer-BioNTech Fall Campaign, and No Vaccine Fall Campaign) during the analysis time horizon between September 1, 2023 to August 31, 2024. The inputs for vaccine effectiveness and coverage are described in the following sections for the base case and scenario analyses.

### Analysis Time Horizon: Fall 2023 COVID-19 Vaccine Effectiveness

For the base case analyses, the updated COVID-19 mRNA Fall 2023 vaccines from both Moderna and Pfizer-BioNTech are assumed to be well-matched to the circulating COVID-19 variant for the entire 1-year time horizon. VE input values are summarized in Table 1. The Moderna updated COVID-19 mRNA Fall 2023 vaccine has been tested in a Phase 2/3 randomized safety and immunogenicity study which demonstrated that it elicits a robust neutralizing antibody response.^21^ As there are no data on clinical outcomes for the Moderna updated Fall 2023 vaccine, data from past vaccine formulations used during the Omicron variant period were assumed. Our estimate of the initial VE against hospitalization was based upon a Kaiser Permanente prospective cohort study of the Moderna bivalent BA.4/BA.5 COVID-19 vaccine by Tseng et al. (2023)^22^. These VE estimates were achieved for the first month following vaccination but then waned with an absolute monthly decrease based on a meta-analysis of the effect of monovalent vaccines against Omicron BA.1/BA.2.^23^ As this study had no outcomes related to infection, the estimate of the initial VE against infection was based on a meta-analysis of monovalent versions of the vaccines administered during the Omicron period.^24^

**Table 1.** Vaccine effectiveness parameters for the updated COVID-19 mRNA Fall 2023 vaccines.

|  | Infection (%) |  | Hospitalizations (%) |  |
| --- | --- | --- | --- | --- |
|  | Initial VE | Monthly Waning | Initial VE | Monthly Waning |
| Moderna updated COVID-19 mRNA Fall 2023 vaccine |  |  |  |  |
| Base case | 57.12% | 4.75% | 84.3% | 1.37% |
| 95% CI for scenario analyses | 30.57%-83.68% | 3.05%-6.75% | 80.3%-87.5% | 0.62%-2.38% |
| Pfizer updated COVID-19 mRNA Fall 2023 vaccine |  |  |  |  |
| Base case | 54.8% | 4.75% | 82.6% | 1.37% |
| Ranges for scenario analyses | 52.0%-55.7% | -- | 81.85% - 83.9% | -- |

VE: vaccine effectiveness; CI: Confidence interval

For the comparison between the Moderna Fall Campaign and the No Fall Vaccine Campaign, several scenario analyses were conducted by varying the initial VE and monthly waning rates according to the 95% confidence intervals (CIs) displayed in Table 1. The VE estimates over time for the base case and scenario analyses are shown in Figure 2.

**Figure 2.**
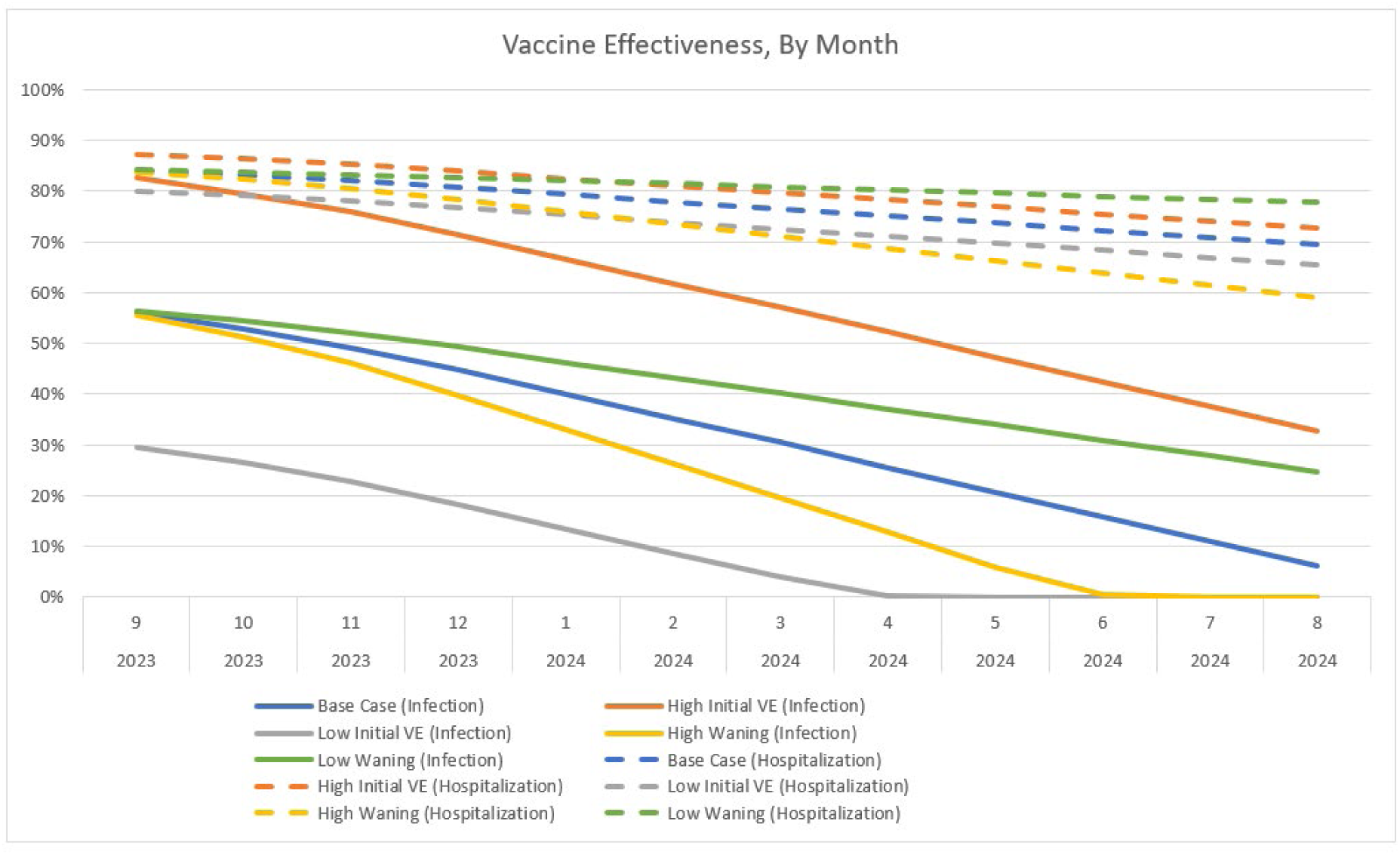
The vaccine effectiveness of the Moderna updated COVID-19 mRNA Fall. 2023 **vaccine over time for those who receive their vaccine in September**.

For the comparison of the Moderna and Pfizer-BioNTech updated COVID-19 mRNA vaccines, the VE of the Moderna vaccine was held constant at the base case value. The relative vaccine effectiveness (rVE) estimates between the two updated Fall 2023 vaccines were assumed based on a US-based comparative retrospective database analysis on the VE of Moderna and Pfizer BA.4/BA.5 bivalent vaccines by Kopel et al. (2023).^18^ For those aged ≥18 years, the rVE for Moderna compared to Pfizer against hospitalizations was 9.8% (95% CI 2.6%-16.4%), while the rVE against outpatient visits, which was used as a proxy for infections, was 5.1% (95% CI: 3.2%-6.9%). For all analyses, the Pfizer VE was calculated based on Moderna VE, reduced by these rVE values, as shown in Table 1. The 95% confidence intervals of the rVEs were used to calculate ranges for scenario analyses. The rVE used for the scenario were only those 65 years and older received the vaccine are presented in the Technical Appendix. The monthly waning rates were assumed to be the same for both the Moderna and Pfizer-BioNTech vaccines in all scenarios.

### Analysis Time Horizon: Fall 2023 COVID-19 Vaccine Coverage

For the base case analyses, uptake of the updated COVID-19 Fall 2023 vaccines was assumed to occur between September 2023 and January 2024, with final coverage rates, by age group, based on the uptake of influenza vaccines for the 2022-23 season.^25^ We assume that individuals who have received at least primary series vaccination have an equal chance of being vaccinated irrespective of their booster histories. Uptake patterns were assumed to be the same for both the Moderna and the Pfizer-BioNTech fall campaigns. For the comparison of the Moderna Fall Campaign to the No Fall Vaccine Campaign, scenario analyses were conducted by varying the uptake rate of those who receive the updated vaccine. Further detail is provided in the Technical Appendix.

### Consequences of Infection

Each run of the SEIR model predicts the total number of infections (asymptomatic and symptomatic) and the incremental reduction in the risk of hospitalization given infection, for vaccinated versus unvaccinated persons for each month of the analysis time period. These monthly outputs are used in a decision tree to calculate the clinical and economic consequences of infections.

The number of monthly infections entering the decision tree is reduced because only symptomatic infections have consequences for quality-of-life and economic costs. For the base case, the proportion of infections that are symptomatic was set to 67.6%^26^ as estimated in a meta-analysis of studies published on Omicron infections. In sensitivity analyses, this proportion was varied according to the 95% CI (60.5% – 74.7%).

Following all symptomatic infections, there is a risk of myocarditis that varies with age. Independent of the risk of myocarditis, a portion of infections are treated in hospital, and the risk of this more severe type of COVID-19 increases with age. The probability of hospitalization in unvaccinated symptomatic patients is displayed in Table 2. For vaccinated persons, these probabilities are reduced according to VE which varies by vaccination strata and which is calculated within the SEIR model. Those who are hospitalized face a risk of mortality that varies by the highest level of care received (ventilator; intensive care unit (ICU) with no ventilator; no ICU or ventilator). There is an increased risk of mortality post-discharge, compared to age-specific mortality rates, as well as a risk of readmission to hospital. The patients who are not hospitalized are considered to be non-severe and are not at risk of death from COVID-19. The inputs for the infection consequences decision tree model are displayed in Table 2. For the DSAs, percentage with symptoms, infection induced myocarditis rates, hospitalization rates in the unvaccinated, hospitalization level of care, and in-hospital mortality rates were varied according to their 95% CI. All other inputs were varied by +/-25% of the base case value.

**Table 2.**
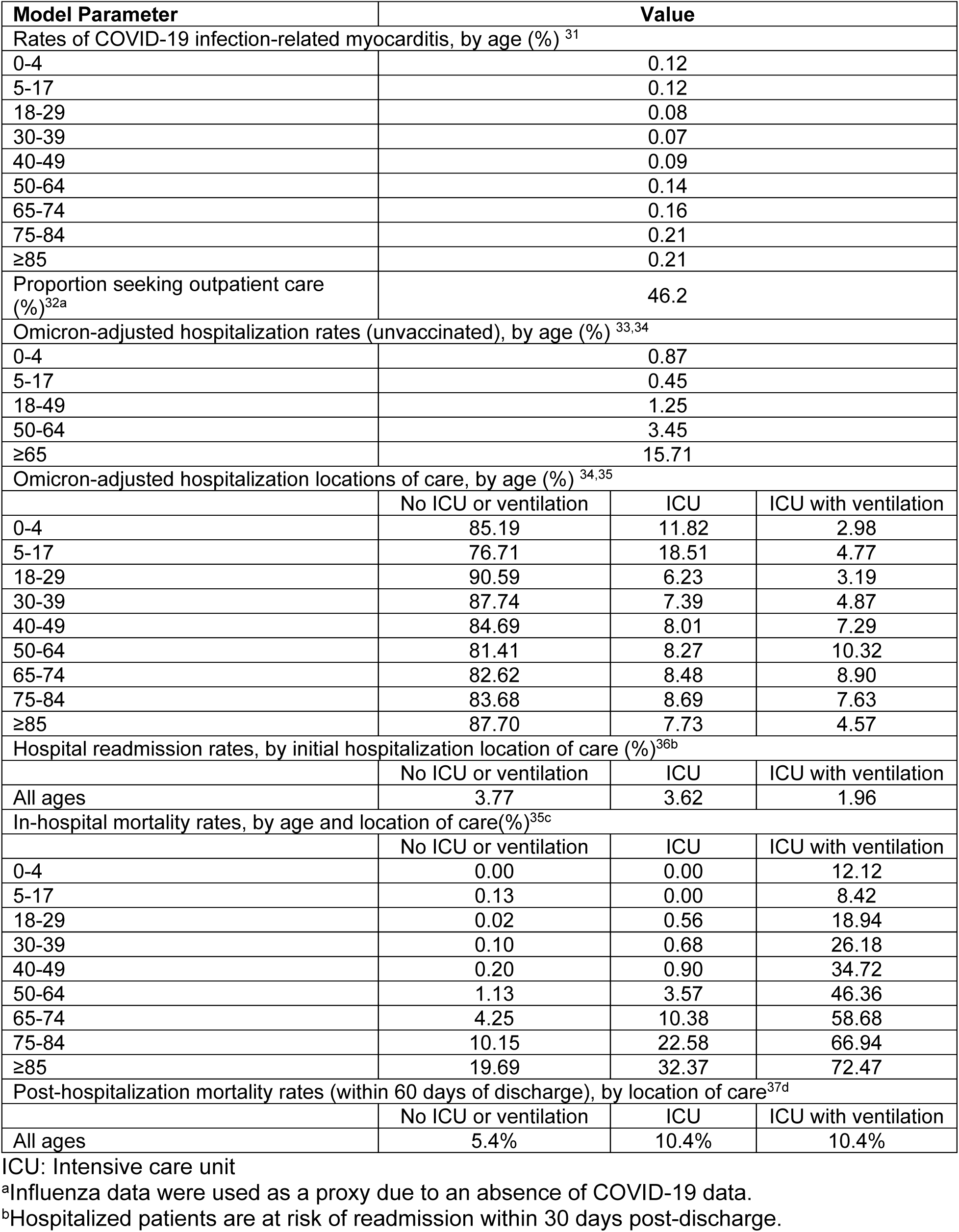
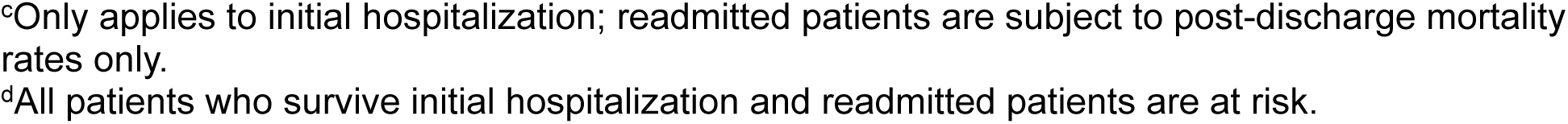
Base case inputs for the infection consequences model.

| Model Parameter | Value |  |  |
| --- | --- | --- | --- |
| Rates of COVID-19 infection-related myocarditis, by age (%) <sup>31</sup> |  |  |  |
| 0-4 | 0.12 |  |  |
| 5-17 | 0.12 |  |  |
| 18-29 | 0.08 |  |  |
| 30-39 | 0.07 |  |  |
| 40-49 | 0.09 |  |  |
| 50-64 | 0.14 |  |  |
| 65-74 | 0.16 |  |  |
| 75-84 | 0.21 |  |  |
| ≥85 | 0.21 |  |  |
| Proportion seeking outpatient care (%)<sup>32a</sup> | 46.2 |  |  |
| Omicron-adjusted hospitalization rates (unvaccinated), by age (%) <sup>33,34</sup> |  |  |  |
| 0-4 | 0.87 |  |  |
| 5-17 | 0.45 |  |  |
| 18-49 | 1.25 |  |  |
| 50-64 | 3.45 |  |  |
| ≥65 | 15.71 |  |  |
| Omicron-adjusted hospitalization locations of care, by age (%) <sup>34,35</sup> |  |  |  |
|  | No ICU or ventilation | ICU | ICU with ventilation |
| 0-4 | 85.19 | 11.82 | 2.98 |
| 5-17 | 76.71 | 18.51 | 4.77 |
| 18-29 | 90.59 | 6.23 | 3.19 |
| 30-39 | 87.74 | 7.39 | 4.87 |
| 40-49 | 84.69 | 8.01 | 7.29 |
| 50-64 | 81.41 | 8.27 | 10.32 |
| 65-74 | 82.62 | 8.48 | 8.90 |
| 75-84 | 83.68 | 8.69 | 7.63 |
| ≥85 | 87.70 | 7.73 | 4.57 |
| Hospital readmission rates, by initial hospitalization location of care (%)<sup>36b</sup> |  |  |  |
|  | No ICU or ventilation | ICU | ICU with ventilation |
| All ages | 3.77 | 3.62 | 1.96 |
| In-hospital mortality rates, by age and location of care(%)<sup>35c</sup> |  |  |  |
|  | No ICU or ventilation | ICU | ICU with ventilation |
| 0-4 | 0.00 | 0.00 | 12.12 |
| 5-17 | 0.13 | 0.00 | 8.42 |
| 18-29 | 0.02 | 0.56 | 18.94 |
| 30-39 | 0.10 | 0.68 | 26.18 |
| 40-49 | 0.20 | 0.90 | 34.72 |
| 50-64 | 1.13 | 3.57 | 46.36 |
| 65-74 | 4.25 | 10.38 | 58.68 |
| 75-84 | 10.15 | 22.58 | 66.94 |
| ≥85 | 19.69 | 32.37 | 72.47 |
| Post-hospitalization mortality rates (within 60 days of discharge), by location of care<sup>37d</sup> |  |  |  |
|  | No ICU or ventilation | ICU | ICU with ventilation |
| All ages | 5.4% | 10.4% | 10.4% |

ICU: Intensive care unit
<sup>a</sup>Influenza data were used as a proxy due to an absence of COVID-19 data.
<sup>b</sup>Hospitalized patients are at risk of readmission within 30 days post-discharge.
<sup>c</sup>Only applies to initial hospitalization; readmitted patients are subject to post-discharge mortality rates only.
<sup>d</sup>All patients who survive initial hospitalization and readmitted patients are at risk.

The base-case economic analyses were conducted using the healthcare perspective, and all cost inputs are shown in Table 2. A scenario analysis was conducted using the societal perspective, which also included lost productivity costs (See Appendix Table 8). All past costs were inflated to 2022 US dollars using the medical care component of the US Consumer Price Index.^27^ For the health care cost perspective, the average cost of ambulatory care was calculated as a mix of treatment in outpatient clinics or the emergency room. As a portion of patients with non-severe COVID-19 will not require any medical attention, the cost of ambulatory care was weighted by the probability of seeking outpatient care for treatment. All patients who recovered from their infection were assigned a post-infection cost which was higher for those treated in hospital compared to those with non-severe cases. As the time horizon for the analytic period is only 1 year, none of the costs were discounted.

The expected number of life years lost due to early deaths from COVID-19 was calculated using expected survival by age as reported by the National Center for Health Statistics.^28^ Age-specific utility values for individuals without infection, obtained using the EuroQol-5D (EQ-5D) in a group of US adults, were attached to each year lost due to early death from COVID-19.^29^ All future QALYs lost were discounted by 3% annually to present value.^30^ The QALYs lost due to morbidity associated with COVID-19 include myocarditis due to COVID-19 infection, COVID-19 treatment, and post-infection care and symptoms (regardless of whether or not medical care was sought), are shown in Table 3. The QALYs lost due to mortality, morbidity and adverse events following vaccination (described below) were summed to determine the total QALYs lost.

**Table 3.**
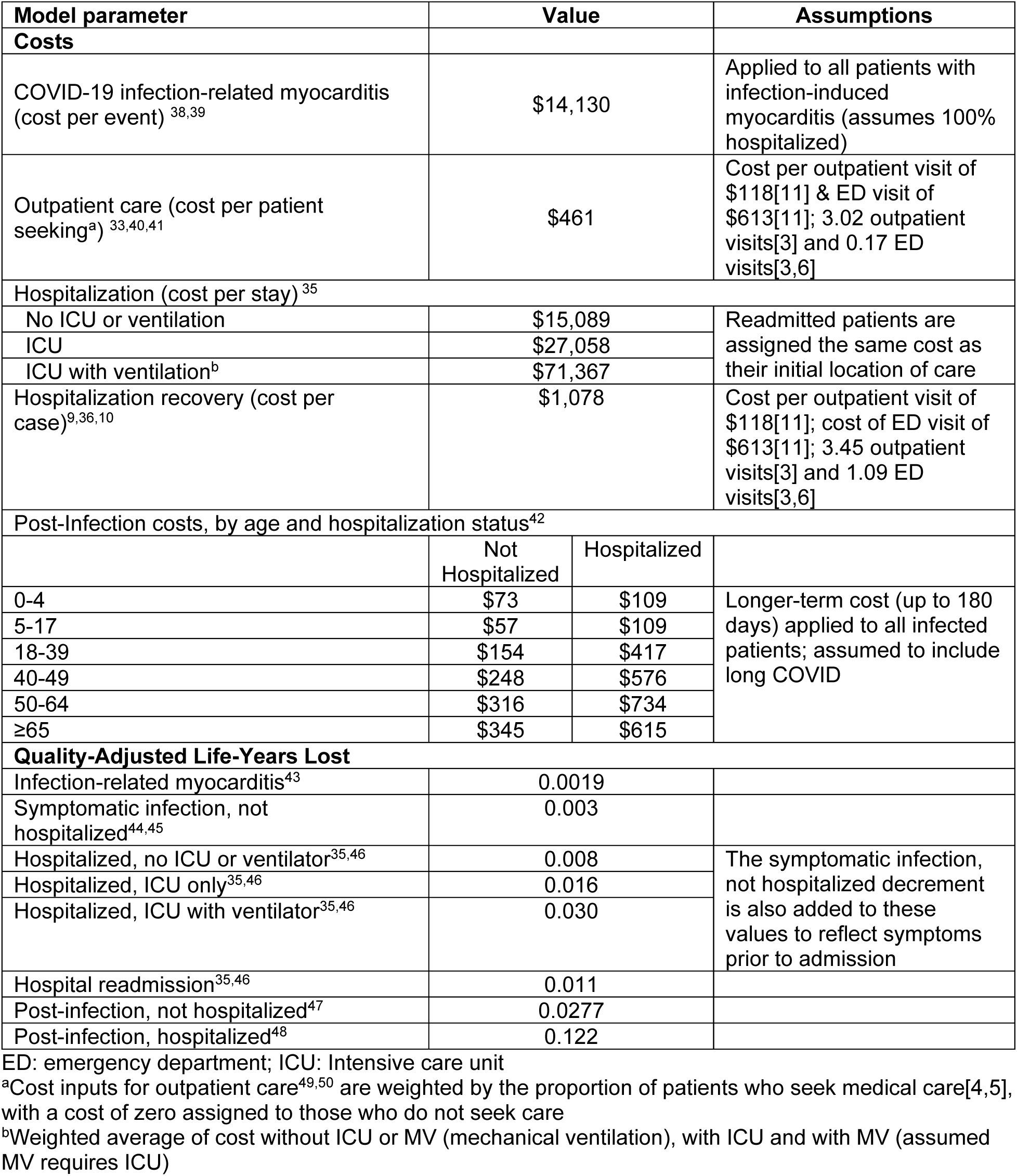
Base case inputs for the infection consequences model (costs and quality-adjusted life-years (QALYs) lost)

### Vaccine Costs and Adverse Events

For each vaccine administered, individuals were assigned a probability of experiencing a vaccine-related adverse event (Grade 3 local or systemic infection-related events; anaphylaxis; myocarditis/pericarditis). The probabilities, along with the associated costs and QALYs lost are displayed in Appendix Table 9. The unit cost of the Moderna updated Fall 2023 vaccine was set to $129.50,^51^ and the unit cost of the Pfizer-BioNTech updated vaccine was assumed to be the same in the base case analysis. A vaccine administration cost of $20.33 was also included in model analyses.^52^ For the societal perspective, time loss from vaccination was assumed to be 0.04 days, considering the majority of COVID-19 vaccine are administered in the pharmacy setting.^16^

### Deterministic Sensitivity Analyses

A series of one-way and multi-way deterministic sensitivity analyses (DSAs) were conducted to test the robustness of results for the comparison of the Moderna Fall campaign to No Vaccine Fall campaign. As described throughout the text and summarized in Appendix Table 11, inputs related to incidence of infection, Fall 2023 vaccine coverage, Fall 2023 vaccine VE, percent of symptomatic infections, and the inputs to the consequences decision tree were varied. A scenario analysis was conducted for both of these comparisons where the target population was limited to vaccination of those aged ≥65 years, rather to than all adults aged ≥18 years. A second scenario analyses was conducted with the societal perspective.

We also evaluated the price difference between the two vaccines that would be economically justifiable (i.e., would make each vaccine the cost-effective choice) at different thresholds of willingness-to-pay (WTP) for QALYs, under different relative VE (rVE) assumptions. The base case model with the healthcare cost perspective was used. As the price of the Pfizer-BioNTech vaccine is not yet known, the price of the Pfizer-BioNTech vaccine remained at $129.50 as in the base case. As the Moderna Fall 2023 vaccine was assumed to be more effective than the Pfizer-BioNTech Fall 2023 vaccine based on comparative data from past formulations, it would dominate the Pfizer-BioNTech vaccine if it was priced the same or lower. Therefore, the unit price of the Moderna vaccine was varied upwards for this analysis.

### Probabilistic Sensitivity Analyses

For the No Vaccine Fall Campaign versus the Moderna Fall Campaign, the percent of patients with symptomatic infections, as well as the probabilities, costs and QALY inputs in Tables 2, 3 and Appendix Table 9, were varied in a probabilistic sensitivity analysis (PSA). Distributions for the PSA were chosen based on recommendations from Briggs, Sculpher, and Claxton.^53^ For skewed data (e.g., costs), for example, a gamma or log-normal distribution is advised, while for binomial data, a beta distribution was used. If data were not available to estimate a standard error, then it was assumed to be 10% of the mean.

## Results

### Comparison: Moderna updated COVID-19 mRNA Fall 2023 vaccine versus No Fall 2023 Vaccine

For the base case scenario, the model predicted that from September 1^st^, 2023 until August 31^st^, 2024 there will be 64,203,464 symptomatic infections if no Fall 2023 COVID-19 vaccines are administered and 56,956,517 if the Moderna updated COVID-19 mRNA vaccine is administered. This represents a decrease of 7,237,947 symptomatic infections (–11%) when 111,097,782 vaccines are administered to those aged 18 years and older. The predicted monthly incidence per 100,000 persons is shown over time, by age group, in Figure 3. Without the vaccine, the incidence is expected to peak in December 2023 and decline to a low point from July to August 2024. With the vaccine, the peak is flattened and incidence is predicted to remain more constant over time. In July and August 2024, the incidence is higher with vaccine compared to no vaccine. This pattern is predicted because the vaccine protection against infection wanes, leading to a rebound in the risk of transmission with contacts at the end of the 1-year time horizon.

**Figure 3.**
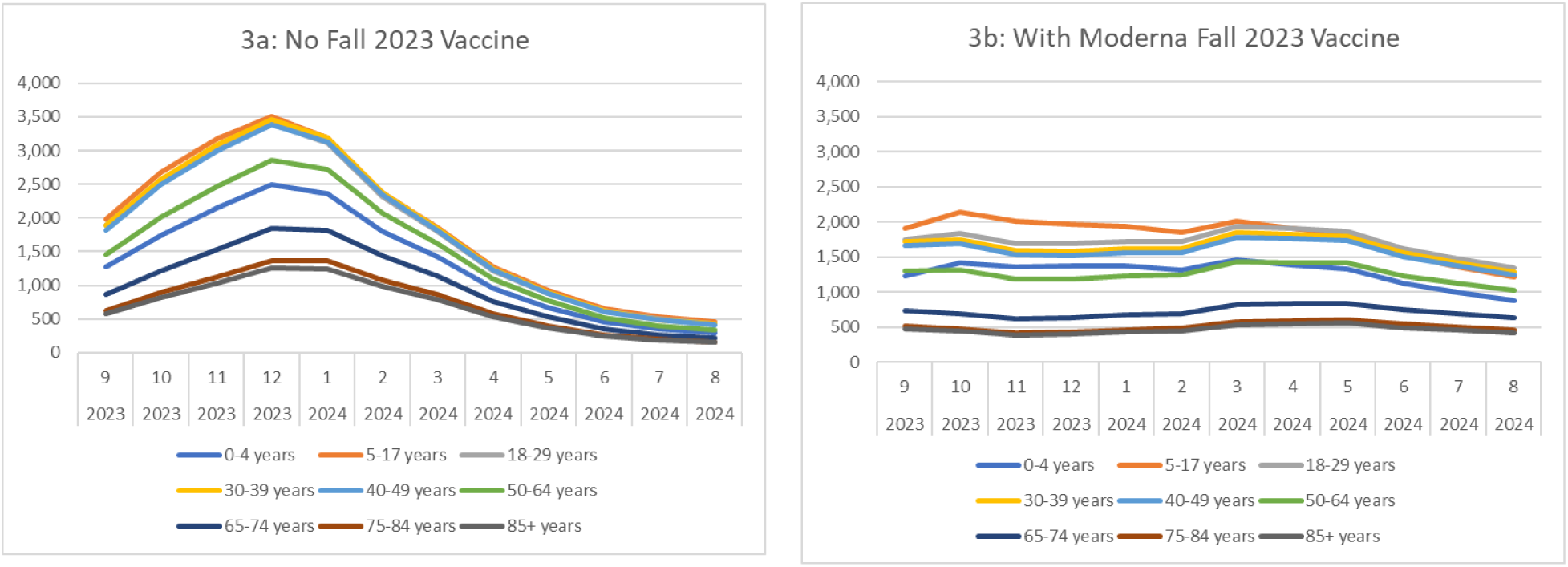
Monthly rate of symptomatic infections (per 100,000 persons) with and without a Moderna updated mRNA Fall 2023 vaccination, by age group.

Figure 3 also illustrates that incidence of symptomatic infection is highest in the 5 to 17 years age group and lowest in those ages 75 years and above. With the No Vaccine Fall Campaign, the residual VE from past vaccination (not shown in the figure) is low, ranging from 4% to 8% among those who received the bivalent booster, to 0% among people who were vaccinated with any number of monovalent doses. The proportion who are immune to infections is higher in those under 50 years of age (ranging from 50.8% to 67.5%) but is 30.9% or lower in the older age groups, suggesting that the incidence patterns by age group are driven by contact patterns and mixing across age groups. With a Fall 2023 vaccine strategy targeting those aged ≥18 years, the 5 to 17 years age group continues to have the highest incidence while ≥75 years has the lowest.

In the base case analysis, with the No Vaccine Fall Campaign, there are 1,199,429 COVID-19 related hospitalizations and 154,798 associated deaths predicted from September 2023 to August 2024. With a Moderna updated mRNA Fall 2023 vaccine, those numbers decline to 856,823 (–29%) and 104,304 (–33%) respectively. The residual VE against hospitalization is higher compared to infection, ranging by age group from 31% to 37% among those with primary series only and 68% to 70% among those with the bivalent booster. While individuals aged ≥65 years have the highest vaccine coverage rates overall, they also have the highest risk of hospitalization and death. As displayed in Figure 4, they are therefore expected to have the highest annual rates of hospitalization and death per 100,000 persons with and without the Fall 2023 vaccine.

**Figure 4.**
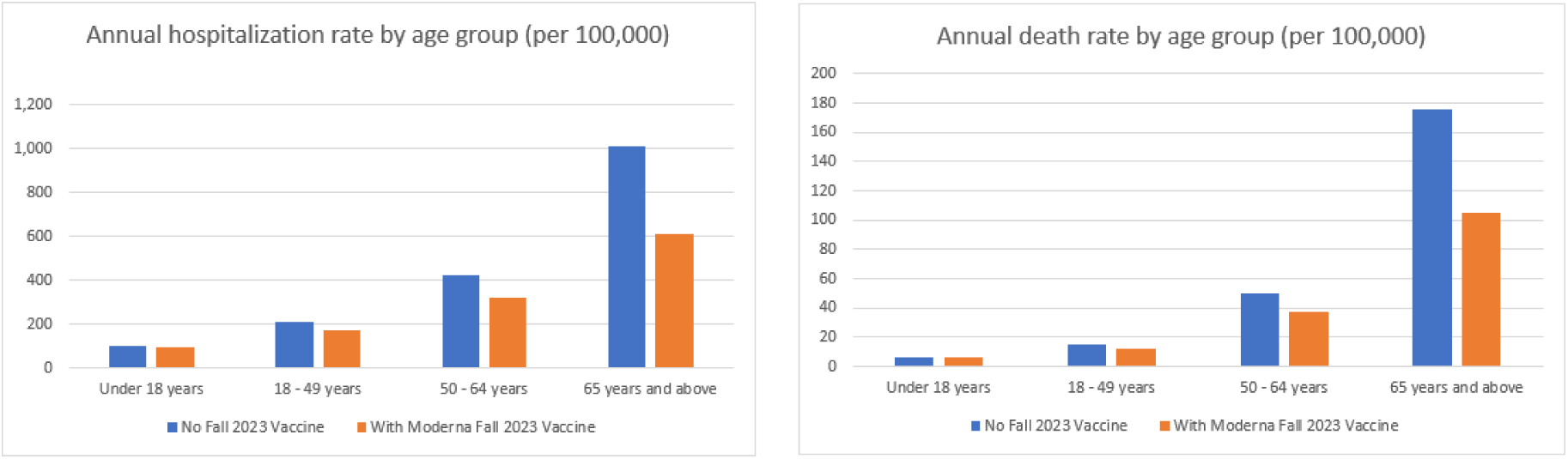
The annual rate of hospitalizations and deaths per 100,000 with and without a Moderna updated mRNA Fall 2023 vaccination across the 1-year time horizon, by age group.

Given this clinical impact, the Moderna updated COVID-19 mRNA Fall 2023 vaccine is expected to lead to a gain of 499,660 QALYs by preventing COVID-19-related deaths and 241,210 QALYs gained due to prevented morbidity for a total of 740,880 QALYs gained (0.007 QALYs gained per vaccination). Approximately 94% of those QALYs gained due to morbidity are due to the prevention of the post-infection impact of COVID-19, while only 8% are from preventing the initial infection itself. Vaccination costs $16,646 million, however, COVID-19 treatment costs with vaccination are $42,330 million compared to $53,255 million without the vaccine, leading to $10,924 million in health care treatment costs prevented. The incremental cost per QALY gained of the Moderna updated COVID-19 mRNA Fall 2023 vaccine compared to no additional COVID-19 vaccination in Fall 2023 is therefore $7,700 (Table 4). See Appendix Table 10 for further cost disaggregation.

**Table 4.** Economic results for the base case and scenario analyses.

| **Vaccination Strategy** | **Total Costs** | **Total QALYs Lost** | **Δ Costs** | **Δ QALY Gained\*** | **ICER (Δ Cost/QALY Gained)** |
| --- | --- | --- | --- | --- | --- |
| Base-case: No additional COVID-19 vaccination in Fall 2023 vs. Moderna updated Fall 2023 vaccine |  |  |  |  |  |
| No Vaccine | \$53,254,666,452 | 3,875,029 | -- | -- | Ref |
| Moderna | \$58,976,187,224 | 3,134,151 | \$5,721,520,772 | 740,878 | \$7,723 |
| Scenario analysis: Adults ≥65 years |  |  |  |  |  |
| No Vaccine | \$53,254,666,452 | 3,875,029 | -- | -- | Ref |
| Moderna | \$53,878,872,864 | 3,532,532 | \$624,206,412 | 342,497 | \$1,823 |
| Scenario analysis: Societal perspective |  |  |  |  |  |
| No Vaccine | \$92,047,025,703 | 3,875,029 | -- | -- | Ref |
| Moderna | \$93,582,793,851 | 3,134,151 | \$1,535,768,148 | 740,878 | \$2,073 |
| Comparison: Pfizer-BioNTech updated Fall 2023 vaccine vs. Moderna updated Fall 2023 vaccine |  |  |  |  |  |
| Moderna | \$58,976,187,224 | 3,134,151 | -- | -- | -- |
| Pfizer-BioNTech** | \$59,917,727,937 | 3,199,569 | \$941,540.713 | 65,418 | Moderna vaccine dominates Pfizer-BioNTech vaccine |

Δ Difference; ICER – incremental cost effectiveness ratio; QALY – quality-adjusted life-year
\* Difference in QALYs gained is equivalent to negative one times the difference in total QALYs lost
\*\*Assuming same cost for vaccine unit and administration fees

When the target population for vaccination was limited to those 65 years and older only, fewer vaccines were delivered (39,793,511) and only 2.7 million cases of symptomatic disease, 180,000 hospitalizations and 31,000 deaths were prevented compared to the 7.2 million cases, 343,000 hospitalizations and 50,000 deaths prevented when all adults are targeted. Older age groups are at greater risk of severe outcomes, so the incremental cost per QALY gained decreases to $1,800 (Table 4). When vaccine coverage rates are decreased, fewer clinical events are prevented (4.0 million symptomatic infections, 181,000 hospitalizations, 27,000 deaths) but, because of increased efficiency due to the non-linear herd effects in the dynamic model, the ICER decreases to $6,200. With the bivalent vaccine, overall coverage was lower than the base case for the new vaccine, but the proportion of older (i.e., higher risk) individuals increases and the incremental cost per QALY gained decreases to $3,900. Using the societal cost perspective, the cost per QALY gained is lower than in the health care perspective, at $2,100 per QALY gained (Table 4).

The 12 most influential variables from the DSAs are summarized in the tornado diagram in Figure 5 (full DSA results are also provided in Appendix Table 11). Overall, vaccine effectiveness, patterns of COVID-19 incidence, hospitalization rates or costs have the greatest impact on the cost-effectiveness.

**Figure 5.**
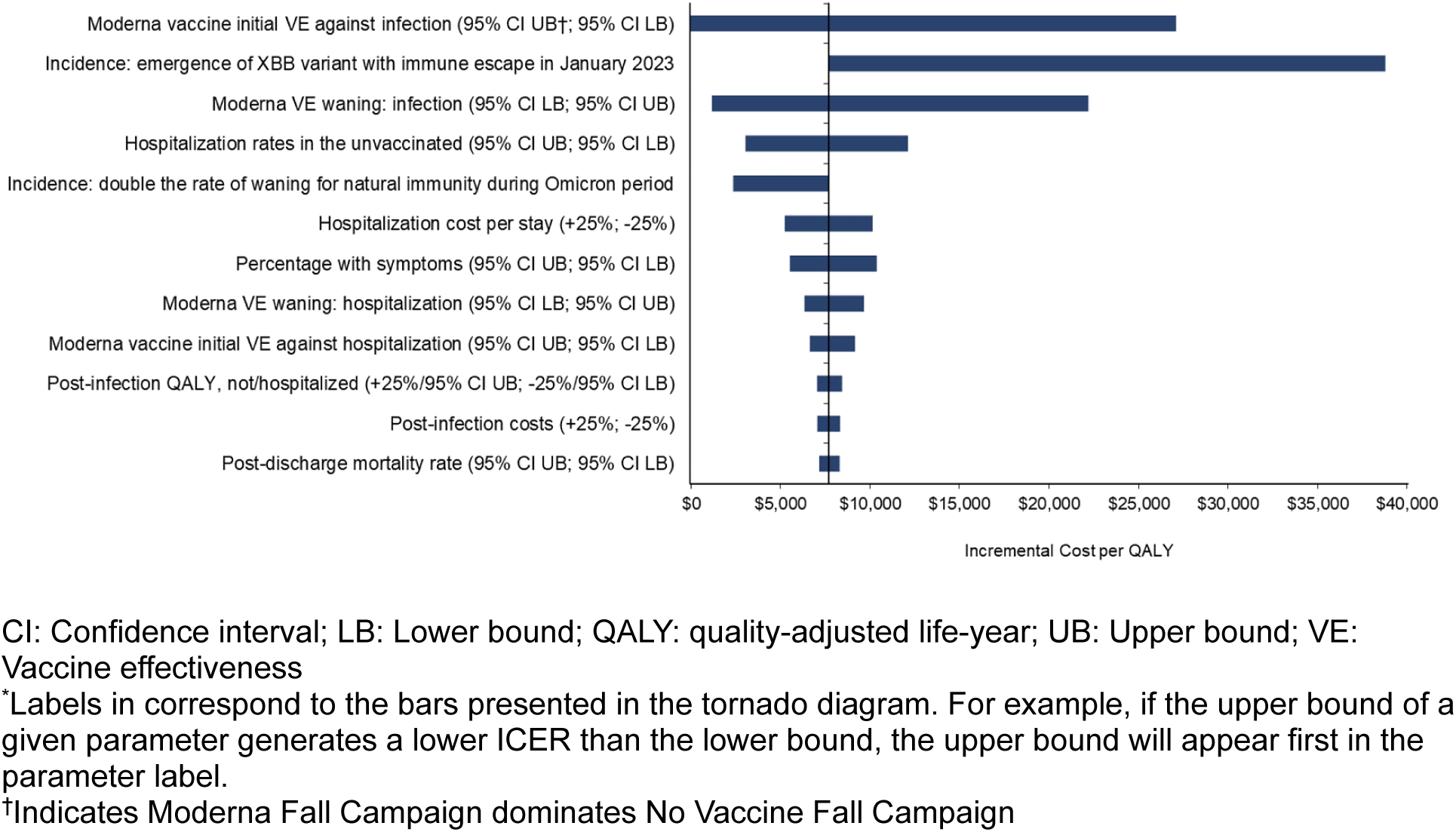
Tornado diagram*: Key deterministic sensitivity analyses.

Varying the Moderna updated COVID-19 mRNA Fall 2023 initial VE against infection has the greatest impact on the ICER. With low VE against infection, the cost per QALY gained increases to $27,100, while at the highest VE the Moderna Fall Campaign becomes cost saving as well as more beneficial compared to no vaccine. If the waning rate of VE against infection is varied, then the ICER ranges from $1,200 to $22,200. Varying the waning rate of VE against hospitalization leads to a smaller range ($6,400 to $9,700). When initial VE against infection and hospitalization are varied simultaneously (not shown in the figure), the ICER ranges from the Moderna Fall Campaign becoming cost saving as well as more beneficial compared to no vaccine to $33,200.

DSA results also suggest that patterns of COVID-19 incidence impact the ICER. Four different incidence scenarios were created with the SEIR model in order to test the potential impact (two of which appear in Figure 5), but the results are complex given the non-linearity of the model. Figure 6 compares the monthly incidence of the base case and the four scenarios. The incidence of infection increases when the rate of waning natural immunity during Omicron is doubled (gray curve). There are more infections, and more prevented infections, with the same number of vaccinations as in the base case, so the ICER drops to $2,400. When the waning rate of natural immunity is halved (green curve), however, the ICER is reduced (improved) slightly to $7,600. While there are fewer infections, the timing of those infections differs, and a greater number are prevented than in the base case. If we consider that the new XBB variant causes a 50% decrease in both vaccine-mediated and natural immunity in the base case, when the model is recalibrated to achieve the same number of infections in the burn-in period the transmissibility parameter is lowered. This in turn, leads to a lower number of infections for all of the analytic period (red curve). In that scenario, there are fewer infections prevented with the vaccine. Therefore, the ICER increases to $38,700. When the percentage of infections that are symptomatic is varied, the cost per QALY ranges from $5,600 to $10,400.

**Figure 6.**
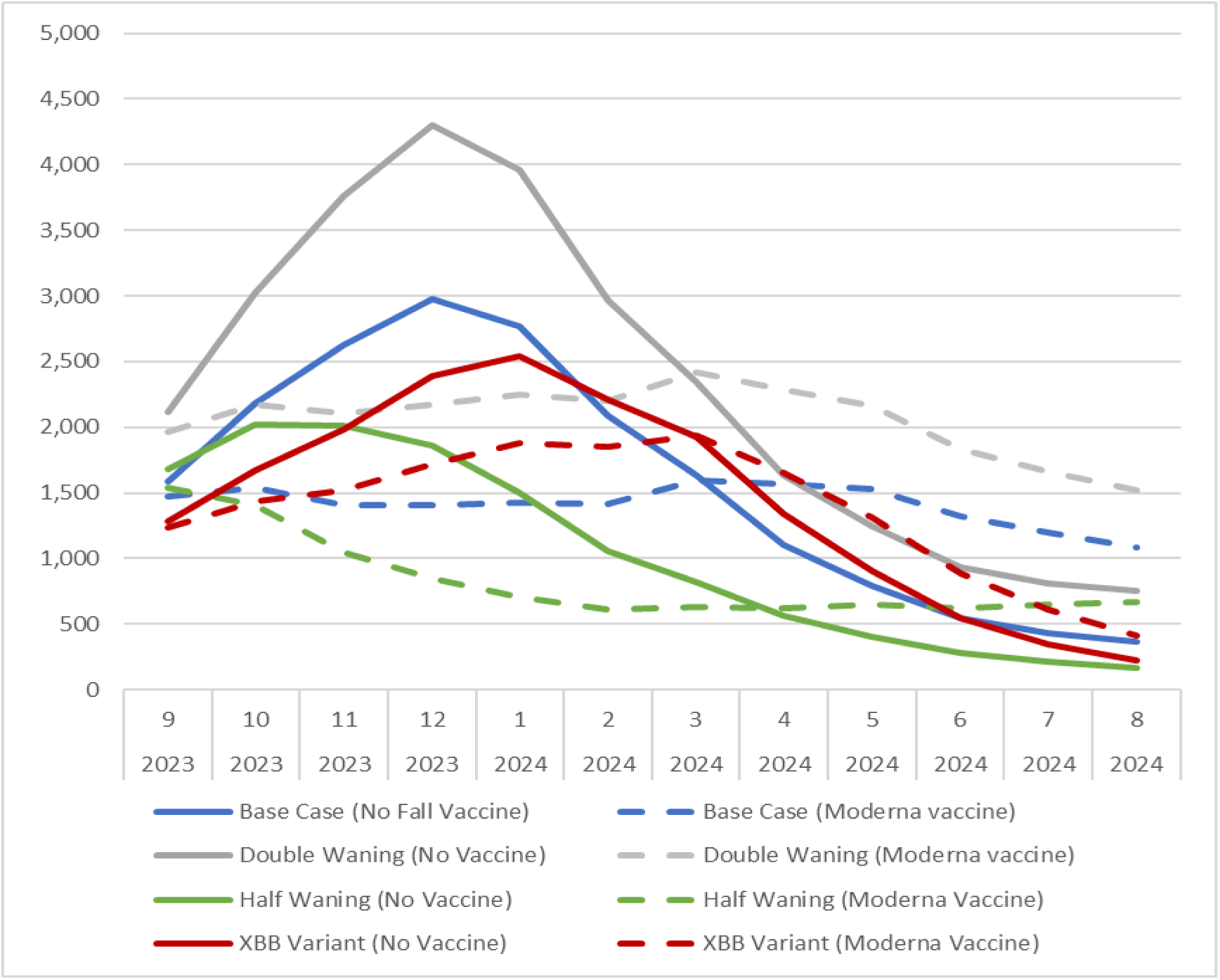
The monthly incidence of symptomatic infection (per 100,000 persons) for base case and three scenario analyses.

When the hospital rate among unvaccinated persons with a symptomatic infection is varied, the incremental cost per QALY gained varied from $3,000 to $12,100. Varying hospital costs by 25% caused the ICER to vary from $5,300 to $10,200. The remaining parameters in the DSA had minimal impact on cost-effectiveness results.

In the probabilistic sensitivity analysis, as shown in the cost-effectiveness acceptability curve in Appendix Figure 11, the Moderna Fall 2023 vaccine reaches a 100% probability of being the most cost-effective option at a willingness-to-pay threshold of approximately $17,000/QALY.

### Comparison: Moderna strategy versus Pfizer strategy

In the base case analysis, the Moderna updated Fall 2023 vaccine had greater initial VE than the Pfizer version of the vaccine. Across the 1-year time frame, the Moderna Fall Vaccine campaign is predicted to prevent 932,165 COVID-19 more infections than the Pfizer Fall Vaccine campaign. In addition, use of the Moderna vaccine is predicted to result in 23,658 fewer hospitalizations and 3,340 fewer deaths than use of the Pfizer-BioNTech vaccine. This is expected to lead to an additional 65,418 QALYs gained. If the vaccines have the same unit cost, then the total costs associated with the use of the Moderna vaccine is expected to be $942 million lower (or $8.47 in treatment costs prevented per vaccination administered) (Table 4). See Appendix Table 10 for further cost disaggregation. With lower total healthcare costs and higher QALYs gained, the Moderna updated COVID-19 mRNA Fall 2023 vaccine dominates the Pfizer-BioNTech one.

The results of the DSA varying the price of the Moderna vaccine are shown in Figure 7 for three different WTP thresholds. As the Moderna vaccine is more effective in our analyses, only the results of where the Moderna is priced at a premium to the Pfizer-BioNTech vaccine are shown. As the rVE of Moderna versus Pfizer-BioNTech increases, the economically justifiable price premium increases. The lines on the graph represent the base case and 95% CI for the rVE data used in this analysis. For the base case rVE (5.10% infection; 9.8% hospitalization), the Moderna Fall Vaccine campaign would be considered as cost-effective compared to the Pfizer-BioNTech Fall Vaccine campaign with a price of $167.42 (price difference of $37.92), $196.86 (price difference of $67.36) and $226.30 (price difference of $95.80) at the WTP thresholds of

**Figure 7.**
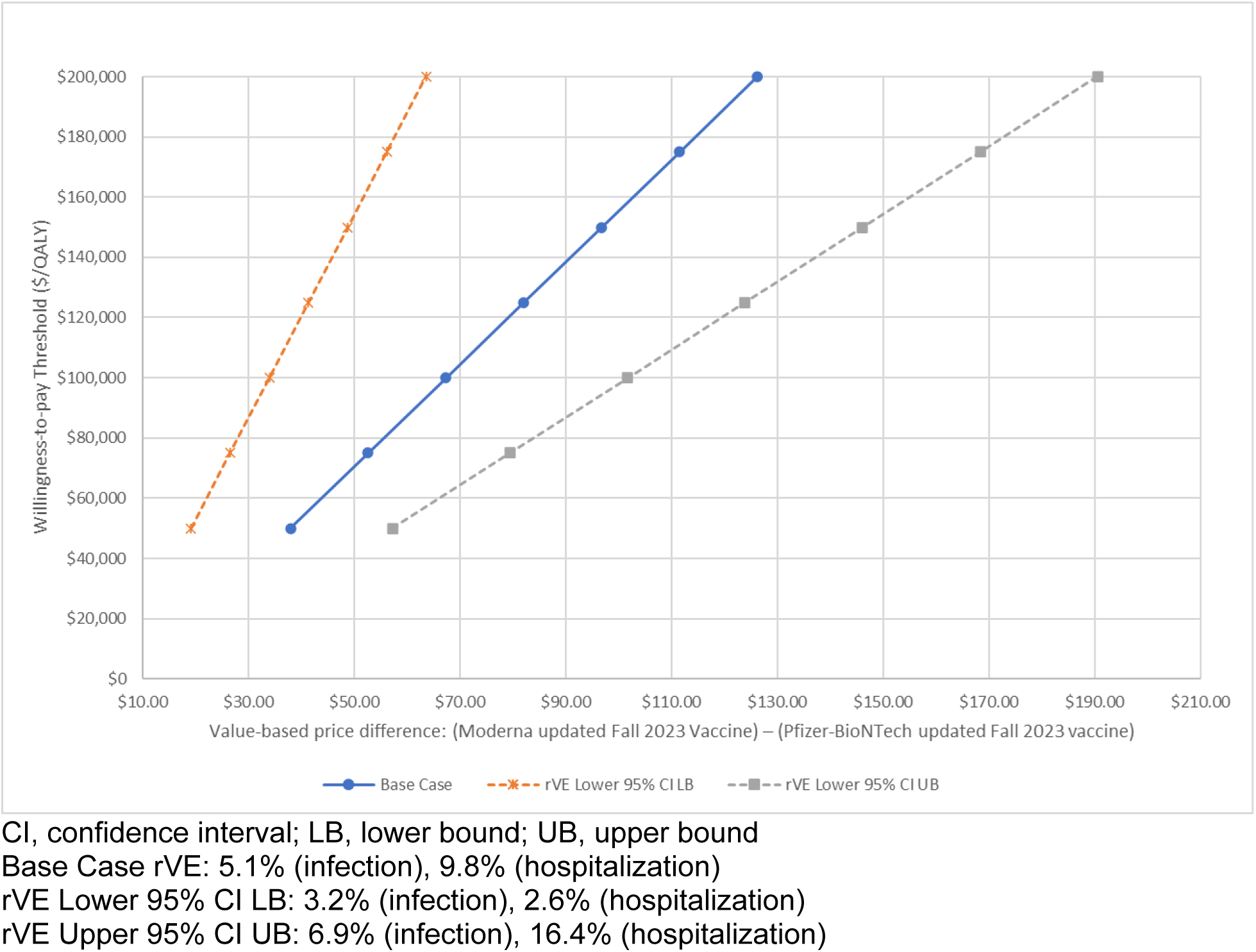
Incremental cost-effectiveness of the Moderna updated Fall 2023 vaccine compared with the Pfizer-BioNTech updated Fall 2023 vaccine across a range of price differences and relative vaccine effectiveness values.

$50,000; $100,000; and $150,000 per QALY gained respectively. As the difference in the VE between the vaccines increases or decrease, the price premium that could be justified at any WTP threshold also increases or decreases.

## Discussion

Using an SEIR model stratified by vaccination status, we examined the potential clinical impact and the cost-effectiveness of Fall 2023 COVID-19 vaccines in the US targeting those 18 years and older. The Moderna updated COVID-19 mRNA Fall 2023 Vaccine is predicted to prevent 7.2 million cases of symptomatic infection, 340,000 hospitalizations and 50,500 deaths between September 2023 and August 2024 compared to no new Fall 2023 vaccination. In the base case analysis, the incremental cost per QALY gained was predicted to be $7,700 for the healthcare payer perspective. Neumann et al. have argued a willingness-to-pay threshold of $100,000 per QALY gained is reasonable for the US.^54^ Considering this threshold, use of the vaccine is cost-effective, or good value for money. Using a societal cost perspective, the ICER was only $2,100 per QALY gained. There is still much uncertainty about the morbidity and mortality associated with COVID-19, but the sensitivity analyses demonstrated that the results are robust. Although the incremental cost per QALY is most affected by the incidence of infection, the VE, and the probability of hospitalization and its associated costs, vaccination with the Moderna updated COVID-19 mRNA Fall 2023 vaccine is cost-effective across a wide range of parameter values and scenarios.

As COVID-19 is transitioning from a pandemic to an endemic state, the future incidence of infection and the expected pattern of infection are highly uncertain. It is clear that the incidence has been declining since the peak of infection in January 2022,^55^ but it is difficult to quantify the change because of the evolution of data collection practices. During the pandemic phase, COVID-19 was reported to public health authorities at a much greater rate than other infectious diseases as many individuals sought confirmatory PCR testing from health care providers.

Since the emergence of Omicron, health care seeking behavior has declined and so has the proportion of infections reported. For this analysis, four different incidence scenarios were generated by the SEIR model to vary the dynamics of infection. For all of these scenarios, use of the Moderna vaccine remained cost-effective as the ICER was below $40,000 per QALY gained.

The VE of the new Fall 2023 vaccines against both hospitalization and infection are unknown. In our analysis, the VE against hospitalization of the bivalent vaccine was used to represent the VE against hospitalization of a Fall 2023 vaccine that is well-matched to the circulating variant.^22^

Data on the VE against infection are more variable, and therefore the results of a meta-analyses were used.^24^ While a poorly matched vaccine would provide some protection, it would likely be associated with lower initial vaccine effectiveness and possibly also faster waning of response. It is also not known when a new variant that evades immunity mediated by past infections (natural immunity) or vaccine doses (vaccine immunity) will emerge. The only option at the point of time when decisions about adopting the new vaccine are made is to examine the impact of a vaccine with a range of VE profiles, as has been done within the scenario analyses. These analyses show that a Fall 2023 COVID-19 vaccine is likely to have good value for money over a range of VE scenarios.

The hospitalization rates in unvaccinated that were used to populate the model were estimated using data from the first year of the pandemic.^33^ They were adjusted downwards in order to account for the lower severity of infections with Omicron variants.^34^ An important driver of hospitalization costs is the level of care required, with individuals in ICU (with or without mechanical ventilation) costing more than other hospitalized patients. The current severity of disease associated with Omicron is not fully quantified and the virulence of future COVID-19 variants is unknown. In sensitivity analyses, however, use of the Fall 2023 vaccine remained cost-effective, with all ICERs falling well below the $100,000 cost per QALY threshold.

Other parameters, which may increase the value of vaccination, were not included in the analysis. For example, although post-infection costs and QALY decrements were included, they were of limited duration. Bowe et al., recently reported on lasting health consequences 2 years post infection.^56^ Zhang et al., found higher rates of newly onset hypertension in post-COVID individuals.^57^ Additionally, although analyses from the societal perspective was included, the emphasis was on short-term lost productivity for the COVID-19 patient. Broader aspects of lost productivity, such as caregiver time,^58,59^ are not included in the current analysis, nor are the consequences of weighing utility losses associated with severe disease higher than non-severe disease and the associated utility losses for families. ^59^ Other parameters, as proposed by the International Society of Pharmacoeconomics and Outcomes Research in the “value flower” framework,^59^ and noted by Postma et al.,^58^ and Di Fusco et al.,^60^ (equity, productivity of other individuals, health system strengthening), which may also increase the value of vaccination, are also not included.

Our analysis quantified the potential clinical and economic impact of using a Fall 2023 COVID-19 vaccine with a higher VE. The Moderna mRNA COVID-19 vaccine was found to be more effective than the Pfizer mRNA COVID-19 vaccine in a meta-analysis in immunocompromised populations^13^ and in studies of the monovalent^14-17^ and bivalent versions^18^ in the general population. While both the Moderna and Pfizer vaccines have the same mechanisms of action, differences in VE may exist because their delivery systems and dosage differ. Assuming both the Moderna and Pfizer-BioNTech vaccines are priced the same, as the Moderna vaccine is predicted to prevent more COVID-19 infections, the net healthcare costs are also predicted to be 941 million lower in the base case. Based on our base case cost-effectiveness analysis, a price premium of up to $67.36 could be economically justifiable with the base case rVE (Moderna versus Pfizer: 5.1% infection; 9.8% hospitalization) considering the $100,000 cost per QALY threshold.

In conclusion, this analysis suggests that using COVID-19 mRNA vaccines in Fall 2023 will still prevent significant morbidity and mortality in US. Furthermore, continued use of the Moderna vaccine is likely to be good value for money as these analyses suggests the vaccine to be highly cost-effective despite the transition from pandemic to endemic.

## Data Availability

All data produced in the present work are contained in the manuscript, and all inputs are fully referenced.

## References

1. United Nations. UN News. WHO chief declares end to COVID-19 as a global health emergency. https://news.un.org/en/story/2023/05/1136367. Updated May 5, 2023.

2. Center for Disease Control and Prevention. End of Federal COVID-19 Public Health Emergnecy (PHE) Declaration. https://www.cdc.gov/coronavirus/2019-ncov/your-health/end-of-phe.html. Updated May 5, 2023.

3. Jones JM, Manrique IM, Stone MS, et al. Estimates of SARS-CoV-2 Seroprevalence and Incidence of Primary SARS-CoV-2 Infections Among Blood Donors, by COVID-19 Vaccination Status – United States, April 2021-September 2022. MMWR Morbidity and mortality weekly report. 2023;72(22):601–605.

4. Centers for Disease Control and Prevention. COVID-19 Vaccine: Interim COVID-19 Immunization Schedule for Persons 6 Months of Age and Older. https://www.cdc.gov/vaccines/covid-19/downloads/COVID-19-immunization-schedule-ages-6months-older.pdf. Published 2023. Accessed June 1, 2023.

5. Hodcroft E. CoVariants. Overview of Variants in Countries: France. https://covariants.org/. Accessed January 23, 2023.

6. Rosenblum HG, Wallace M, Godfrey M, et al. Interim Recommendations from the Advisory Committee on Immunization Practices for the Use of Bivalent Booster Doses of COVID-19 Vaccines – United States, October 2022. MMWR Morbidity and mortality weekly report. 2022;71(45):1436–1441.

7. Link-Gelles R, Ciesla AA, Roper LE, et al. Early Estimates of Bivalent mRNA Booster Dose Vaccine Effectiveness in Preventing Symptomatic SARS-CoV-2 Infection Attributable to Omicron BA.5– and XBB/XBB.1.5-Related Sublineages Among Immunocompetent Adults – Increasing Community Access to Testing Program, United States, December 2022-January 2023. MMWR Morbidity and mortality weekly report. 2023;72(5):119–124.

8. Link-Gelles R, Weber ZA, Reese SE, et al. Estimates of Bivalent mRNA Vaccine Durability in Preventing COVID-19-Associated Hospitalization and Critical Illness Among Adults with and Without Immunocompromising Conditions – VISION Network, September 2022-April 2023. MMWR Morbidity and mortality weekly report. 2023;72(21):579–588.

9. FDA U.S. Food & Drug Administration. Recommendation for the 2023-2024 Formula of COVID-19 vaccins in the U.S. https://www.fda.gov/media/169591/download. Accessed June 16, 2023.

10. Hodcroft E. CoVariants. https://covariants.org/. Accessed January 23, 2023.

11. Utami AM, Rendrayani F, Khoiry QA, et al. Economic evaluation of COVID-19 vaccination: A systematic review. J Glob Health. 2023;13:06001.

12. Kohli MA, Maschio M, Lee A, et al. The potential clinical impact of implementing different COVID-19 boosters in fall 2022 in the United States. J Med Econ. 2022;25(1):1127–1139.

13. Wang X, Haeussler K, Spellman A, et al. Comparative Effectiveness of mRNA-1273 and BNT162b2 COVID-19 Vaccines in Immunocompromised Individuals: A Systematic Review and Meta-Analysis Using the GRADE Framework. medRxiv.2023.04.05.23288195.

14. Nguyen VH BC, Bogdanov A, et al. Relative effectiveness of BNT162b2, mRNA-1273, and Ad26.COV2.S vaccines and homologous boosting in preventing COVID-19 in adults in the US. medRxiv. 2023.

15. Ono S, Michihata N, Yamana H, et al. Comparative Effectiveness of BNT162b2 and mRNA-1273 Booster Dose After BNT162b2 Primary Vaccination Against the Omicron Variants: A Retrospective Cohort Study Using Large-Scale Population-Based Registries in Japan. Clin Infect Dis. 2023;76(1):18–24.

16. Mayr FB TV, Shaikh OS, et al.,. Comparative COVID-19 Vaccine Effectiveness Over Time in Veterans. Open Forum Infect Dis. 2022;9(7).

17. Hulme WJ, Horne EMF, Parker EPK, et al. Comparative effectiveness of BNT162b2 versus mRNA-1273 covid-19 vaccine boosting in England: matched cohort study in OpenSAFELY-TPP. BMJ. 2023;380:e072808.

18. Kopel H, Nguyen VH, Boileau C, et al. Comparative Effectiveness of the Bivalent (Original/Omicron BA.4/BA.5) mRNA COVID-19 Vaccines mRNA-1273.222 and BNT162b2 Bivalent in Adults in the United States. medRxiv. 2023:2023.2007.2012.23292576.

19. Wolfel R, Corman VM, Guggemos W, et al. Virological assessment of hospitalized patients with COVID-2019. Nature. 2020;581(7809):465–469.

20. Imai N, Cori A, Dorigatti I, et al. Report 3: Transmissibility of 2019-nCOV. Imperial College London. (25-01-2020). 10.25561/77148. Published 2020. Updated January 25, 2020. Accessed May 15, 2022.

21. Moderna Inc. Moderna COVID-19 Variant Vaccines. Presentation to the Vaccines and Related Biological Products Advisory Committee. June 15, 2023. https://www.fda.gov/advisory-committees/advisory-committee-calendar/vaccines-and-related-biological-products-advisory-committee-june-15-2023-meeting-announcement#event-materials. Accessed June 23, 2023.

22. Tseng H, Ackerson BK, Sy LS, et al.,. Effectiveness of mRNA-1273 bivalent (Original and Omicron BA.4/BA.5) COVID-19 vaccine in preventing hospitalizations for COVID-19, medically attended SARS-CoV-2 infections, and hospital death in the United States. medRXiv. 2023.

23. Higdon MM, Baidya A, Walter KK, et al. Duration of effectiveness of vaccination against COVID-19 caused by the omicron variant. The Lancet Infectious diseases. 2022;22(8):1114–1116.

24. Pratama NR, Wafa IA, Budi DS, et al. Effectiveness of COVID-19 Vaccines against SARS-CoV-2 Omicron Variant (B.1.1.529): A Systematic Review with Meta-Analysis and Meta-Regression. Vaccines. 2022;10(12):2180.

25. Centers for Disease Control and Prevention. Weekly flu vaccination dashboard: Influenza Vaccination Coverage, Adults. https://www.cdc.gov/flu/fluvaxview/dashboard/vaccination-adult-coverage.html. Published 2023. Accessed May 5, 2023.

26. Shang W, Kang L, Cao G, et al. Percentage of Asymptomatic Infections among SARS-CoV-2 Omicron Variant-Positive Individuals: A Systematic Review and Meta-Analysis. Vaccines (Basel). 2022;10(7).

27. US Bureau of Labor Statistics. US Consumer Price Index: Medical care in US city average, all urban consumers. https://data.bls.gov/cgi-bin/surveymost. Published 2023. Accessed May 15, 2023.

28. Centers for Disease Control and Prevention (CDC). National Center for Health Statistics. National Vital Statistics System – Life Expectancy. https://ftp.cdc.gov/pub/Health_Statistics/NCHS/Publications/NVSR/68_07/. Published 2017. Accessed June 24,, 2020.

29. Hanmer J, Lawrence WF, Anderson JP, Kaplan RM, Fryback DG. Report of nationally representative values for the noninstitutionalized US adult population for 7 health-related quality-of-life scores. Med Decis Making. 2006;26(4):391–400.

30. Sanders GD, Neumann PJ, Basu A, et al. Recommendations for Conduct, Methodological Practices, and Reporting of Cost-effectiveness Analyses: Second Panel on Cost-Effectiveness in Health and Medicine. JAMA. 2016;316(10):1093–1103.

31. Boehmer TK, Kompaniyets L, Lavery AM, et al. Association Between COVID-19 and Myocarditis Using Hospital-Based Administrative Data – United States, March 2020– January 2021. MMWR Morbidity and mortality weekly report. 2021;70(35):1228–1232.

32. Centers for Disease Control and Prevention. Estimated Flu-Related Illnesses, Medical visits, Hospitalizations, and Deaths in the United States — 2019–2020 Flu Season https://www.cdc.gov/flu/about/burden/2019-2020.html. Accessed October 28, 2022.

33. Reese H, Iuliano AD, Patel NN, et al. Estimated Incidence of Coronavirus Disease 2019 (COVID-19) Illness and Hospitalization-United States, February-September 2020. Clin Infect Dis. 2021;72(12):e1010–e1017.

34. Wang L, Berger N, Kaelber D, Davis P, Volkow N, Xu R. COVID infection rates, clinical outcomes, and racial/ethnic and gender disparities before and after Omicron emerged in the US. medRxiv. 2022;2022.02.21.22271300.

35. Di Fusco M, Shea KM, Lin J, et al. Health outcomes and economic burden of hospitalized COVID-19 patients in the United States. J Med Econ. 2021;24(1):308–317.

36. Verna EC, Landis C, Brown RS, Jr., et al. Factors Associated with Readmission in the US Following Hospitalization with COVID-19. Clin Infect Dis. 2021.

37. Chopra V, Flanders SA, O’Malley M, Malani AN, Prescott HC. Sixty-Day Outcomes Among Patients Hospitalized With COVID-19. Ann Intern Med. 2021;174(4):576–578.

38. HCUPnet – Hospital Inpatient National Statistics. 2018 National Diagnoses – Clinical Classification Software Refined (CCSR), Principal Diagnosis: CIR005 Myocarditis and Cardiomyopathy. https://hcupnet.ahrq.gov/#setup. Published 2018. Accessed October 13, 2021.

39. Center for Medicare and Medicaid Services. 2022 National Physician Fee Schedule Relative Value File October Release. https://www.cms.gov/medicare/medicare-fee-for-service-payment/physicianfeesched/pfs-relative-value-files. Published 2022. Accessed May, 2023.

40. Fiedler M, Song Z. Brookings report: estimating potential spending on COVID-19 care. https://www.brookings.edu/research/estimating-potential-spending-on-covid-19-care/. Accessed July 22, 2020.

41. Huang BZ, Creekmur B, Yoo MS, Broder B, Subject C, Sharp AL. Healthcare Utilization Among Patients Diagnosed with COVID-19 in a Large Integrated Health System. J Gen Intern Med. 2022;37(4):830–837.

42. Koumpias AM, Schwartzman D, Fleming O. Long-haul COVID: healthcare utilization and medical expenditures 6 months post-diagnosis. BMC Health Serv Res. 2022;22(1):1010.

43. Prosser LA, Harpaz R, Rose AM, et al. A Cost-Effectiveness Analysis of Vaccination for Prevention of Herpes Zoster and Related Complications: Input for National Recommendations. Ann Intern Med. 2019;170(6):380–388.

44. Smith KJ, Roberts MS. Cost-effectiveness of newer treatment strategies for influenza. The American journal of medicine. 2002;113(4):300–307.

45. Centers for Disease Control and Prevention. Isolation and Precautions for People with COVID-19. https://www.cdc.gov/coronavirus/2019-ncov/your-health/isolation.html. Accessed December 21, 2022.

46. Barbut F, Galperine T, Vanhems P, et al. Quality of life and utility decrement associated with Clostridium difficile infection in a French hospital setting. Health and quality of life outcomes. 2019;17(1):6.

47. Sandmann FG, Tessier E, Lacy J, et al. Long-Term Health-Related Quality of Life in Non-Hospitalized Coronavirus Disease 2019 (COVID-19) Cases With Confirmed Severe Acute Respiratory Syndrome Coronavirus 2 (SARS-CoV-2) Infection in England: Longitudinal Analysis and Cross-Sectional Comparison With Controls. Clin Infect Dis. 2022;75(1):e962–e973.

48. The PHOSP-COVID Collaborative Group. Clinical characteristics with inflammation profiling of long COVID and association with 1-year recovery following hospitalisation in the UK: a prospective observational study. Lancet Respir Med. 2022;10:761–775.

49. Centers for Disease Control and Prevention. Weekly U.S. Influenza Surveillance Report. https://www.cdc.gov/flu/weekly/index.htm. Accessed June 1, 2023.

50. Shao W, Chen X, Zheng C, et al. Effectiveness of COVID-19 vaccines against SARS-CoV-2 variants of concern in real-world: a literature review and meta-analysis. Emerg Microbes Infect. 2022;11(1):2383–2392.

51. IBM Micromedex. Average wholesale price from RedBook NDC accessed via Compendia. Available from: https://www.ibm.com/us-en/marketplace/micromedex-red-book. Accessed August 16, 2023.

52. Centers for Medicare and Medicaid Services. National Physician Fee Schedule Relative Value File April Release. https://www.cms.gov/medicare/medicare-fee-service-payment/physicianfeesched/pfs-relative-value-files/rvu23b. Published 2023. Accessed May 3, 2023.

53. Briggs AH, Schulpher MJ, Claxton K. Decision modelling for health economic evaluation. Oxford, United Kingdon: Oxford University Press; 2006.

54. Neumann PJ, Cohen JT, Weinstein MC. Updating cost-effectiveness--the curious resilience of the $50,000-per-QALY threshold. N Engl J Med. 2014;371(9):796–797.

55. Thornburg NJ. Update on Current Epidemiology of COVID-19 Pandemic and SARS-CoV-2 Variants. Presentation to the Vaccines and Relatd Biological Products Advisory Committee. June 15, 2023. https://www.fda.gov/advisory-committees/advisory-committee-calendar/vaccines-and-related-biological-products-advisory-committee-june-15-2023-meeting-announcement#event-materials. Accessed.

56. Bowe B, Xie Y, Al-Aly Z. Postacute sequelae of COVID-19 at 2 years. Nature medicine. 2023.

57. Zhang V, Fisher M, Hou W, Zhang L, Duong TQ. Incidence of New-Onset Hypertension Post-COVID-19: Comparison With Influenza. Hypertension. 2023.

58. Postma M, Biundo E, Chicoye A, et al. Capturing the value of vaccination within health technology assessment and health economics: Country analysis and priority value concepts. Vaccine. 2022;40(30):3999–4007.

59. Lakdawalla DN, Doshi JA, Garrison LP, Jr., Phelps CE, Basu A, Danzon PM. Defining Elements of Value in Health Care-A Health Economics Approach: An ISPOR Special Task Force Report [3]. Value Health. 2018;21(2):131–139.

60. Di Fusco M MD, Steuten L, et al.,. The Societal Value of Vaccines: Expert-Based Conceptual Framework and Methods Using COVID-19 Vaccines as a Case Study. Vaccines. 2023;11(234).

