## Supplemental Material for "The potential clinical impact and cost-effectiveness of the updated COVID-19 mRNA Fall 2023 vaccines in the United States"

### Technical Appendix

**Tables**

**Figures**

### 1. SEIR MODEL INPUTS: BURN-IN PERIOD (JANUARY 2020 TO AUGUST 2023)

The model structure and underlying assumptions of the Susceptible-Exposed-Infected-Recovered (SEIR) model were described in the Technical Appendix of the previously published analysis.[1]

The probability of infection, or force of infection, is a function of the number of susceptible people in the population as well as the rate of effective contacts between susceptible and infected individuals. Effective contacts are a function of the rate of contact and the transmissibility of the virus per contact. For individuals in the various vaccine strata, the force of infection is reduced based on the average vaccine effectiveness of the cohort.

For the burn-in period (January 2020 to August 2023), the variables that determine the rate of contact and the average vaccine effectiveness were estimated from data as described in sections 1.2 - 1.3 below. The other model transitions (movements into the I, R, and S compartments) were estimated as described in section 1.4. The transmissibility of the virus changed over time as new variants emerged. The transmissibility was therefore estimated through a calibration process described in section 1.5. Any inputs that change in the analytic period are described in section 2.

#### 1.1. Number of Susceptibles

All individuals in the United States were included in the model and were considered to be susceptible to COVID-19 infection at the start of the simulation (January 2020). The size of the US population in 2020 by age was obtained from the United Nations Department of Economic and Social Affairs.[2]

**Table 1. The model population size**

| Age Group (Years) | Number |
|---|---|
| 0-9 | 40,154,632 |
| 10-19 | 43,997,705 |
| 20-29 | 45,113,502 |
| 30-39 | 45,581,347 |
| 40-49 | 41,924,795 |
| 50-59 | 43,489,471 |
| 60-69 | 39,581,587 |
| 70-79 | 23,576,836 |
| 80+ | 12,522,132 |
| **Total** | **335,942,007** |

### 1.2. Number of Effective Contacts

The mixing patterns are based on contact matrices. As behaviors that impacted contact changed during the pandemic, the matrices are modified by a mobility index that accounts for social distancing and mask use.

#### 1.2.1. Mixing Patterns / Contact Matrices

Data on the age-specific mixing patterns in the general population for the United States were obtained from Prem and colleagues.[3] In the published contact matrices, the population was partitioned into 5-year age bands, and all individuals aged 75 years and older were grouped together. For this analysis, the age-specific mixing patterns were first converted into 10-year age bands. Then, we assumed symmetry between the age groups (i.e., an effective contact between someone in age group *i* with someone from age group *j* is the same as an effective contact between someone in age group *j* with someone from age group *i),* weighted by the population estimates in age groups *i* and *j*.

Thus, $c_{ij} = \frac{1}{N_j} \times \frac{\left(c_{ij}^{*} \times N_j\right) + \left(c_{ji}^{*} \times N_i\right)}{2}$ ,

where:

$c_{ij}$ is the number of effective contacts between someone in age group *i* with someone from age group *j*

$c_{ij}^{*}$ is the number of effective contacts between someone in age group *i* with someone from age group *j* based on the original age-specific mixing patterns were first converted into 10-year age bands

$N_i$ is the population size in age group *i*.

The base contact matrix is presented in Table 2.

**Table 2. Base contact matrix used in the model before applying scaling factors**

| Age Group of Participant (Years) | Age Group of Contact (Years) | | | | | | | | |
|---|---|---|---|---|---|---|---|---|---|
| | **0-9** | **10-19** | **20-29** | **30-39** | **40-49** | **50-59** | **60-69** | **70-79** | **80+** |
| **0-9** | 5.0604 | 1.2670 | 0.7715 | 1.7630 | 1.0016 | 0.7916 | 0.5682 | 0.4429 | 0.7981 |
| **10-19** | 1.3333 | 11.7181 | 1.6963 | 1.4153 | 2.1288 | 1.3544 | 0.4907 | 0.6714 | 1.2603 |
| **20-29** | 0.8757 | 1.8296 | 5.6961 | 2.5122 | 2.0989 | 1.7911 | 0.6032 | 0.3591 | 0.5408 |
| **30-39** | 1.9578 | 1.4936 | 2.4581 | 4.3860 | 2.8645 | 1.8419 | 0.9122 | 0.5834 | 0.9352 |
| **40-49** | 1.0153 | 2.0508 | 1.8747 | 2.6148 | 3.7184 | 1.9958 | 0.6981 | 0.8008 | 1.2605 |
| **50-59** | 0.8429 | 1.3706 | 1.6805 | 1.7662 | 2.0966 | 2.8073 | 0.8874 | 0.6151 | 1.0491 |
| **60-69** | 0.5433 | 0.4458 | 0.5081 | 0.7853 | 0.6584 | 0.7967 | 1.4255 | 0.7012 | 0.6706 |
| **70-79** | 0.2637 | 0.3799 | 0.1884 | 0.3128 | 0.4703 | 0.3439 | 0.4367 | 0.8033 | 0.8743 |
| **80+** | 0.2593 | 0.3891 | 0.1548 | 0.2736 | 0.4040 | 0.3201 | 0.2279 | 0.4771 | 0.7925 |

Note: Since the population size of the age groups are not of equal size, the transformed contact matrix is not strictly symmetric.

During the pandemic, the rate of contact was reduced by behaviors such as social distancing and mask use. The magnitude of the impact is estimated as described in the next section.

#### 1.2.2. Social Distancing and Mask Use: Overall Scaling Factor

From January 30, 2020 to June 24, 2022, it was assumed that regular patterns of interaction were modified because of social distancing and use of masks to reduce effective contacts between individuals. Data on social distancing patterns and mask use were obtained from the Institute for Health Metrics and Evaluation (IHME) and used to adjust the base contact matrix based on these factors[4]. Daily estimates on social distancing patterns and mask use were obtained from the IHME for the time period February 2020 through June 24, 2022, inclusive. For the time period after July 31, 2022, a single seasonality parameter was assumed to replace the social distancing patterns and mask use data. Between these two time points (June 24, 2022 and July 31, 2022), a linear interpolation between the scaling factors estimates was assumed to avoid an abrupt transition between these two methods.

For the base case, we assumed that mask use was 0% from July 31, 2022 onwards. We also assumed that social mobility would return to normal (baseline) after this date and stay at this level for the remaining time period.

Mask use represents the percentage of the population who say they always wear a mask in public. Daily estimates of mask use were obtained from IHME[4]. It was assumed that 100% mask usage is associated with a 30% reduction in transmission and that a reduction in mask usage impacts the reduction in transmission proportionately[4]. Therefore, the following scaling factor (due to mask use) was applied daily to the base contact matrix:

$$Scaling\ Factor_{Mask\ use} = 1 - (Mask\ use(\%) \times 0.30)$$

Daily estimates of the change in mobility were obtained from IHME[4] and was applied to reduce the number of contacts per person. Since age-specific data on changes in mobility were not available, the impact was applied equally to all age groups. Therefore, the following scaling factor (due to change in mobility) was applied daily to the base contact matrix:

$$Scaling\ Factor_{Mobility} = 1 + Change\ in\ mobility\ (\%)$$

After July 31, 2022, a standard sinusoidal function was applied to vary the rate of contact by season.[5] The assumed peak was February 15 to account for greater time spent indoors in the winter. The assumed trough was August 15 to account for greater time spent outdoors in summer. The trough and peak were 95% and 105% of normal contact respectively.[6]

We define the seasonality function over time applied to the transmissibility parameter that the transition rate is a function of time $\beta(t)$ as follows:

$$Seasonality = \left(1 + \frac{\phi}{2}\sin(2\pi t + \omega)\right)$$

where $\phi$ denotes the amplitude of seasonality (0.1 in the base case)

and $\omega$ is the phase shift of the sine function such that the peak was February 15 and the trough was August 15.

The overall scaling factor is thus calculated as:

| | | |
|---|---|---|
| Overall Scaling Factor | $[Scaling\ Factor_{Mask\ use}] \times [Scaling\ Factor_{Mobility}]$ | from January 30, 2020 to June 24, 2022 |
| | Seasonality estimate | after July 31, 2022 |
| | Linear interpolation between above | Between June 24 and July 31, 2022 |

| | | |
|---|---|---|
| Overall Scaling Factor | $[1-(Mask\ use(\%)\times 0.30)] \times [1 + Change\ in\ mobility\ (\%)]$ | from January 30, 2020 to June 24, 2022 |
| | $\left(1+\frac{\phi}{2}\sin(2\pi t+\omega)\right)$ | after July 31, 2022 |
| | Linear interpolation between above | Between June 24 and July 31, 2022 |

The final monthly pattern for the scaling factor for both the burn-in period (January 2020 – July 2023) and the analysis period (September 2023 – August 2024) is shown in Figure 1.

**Figure 1. Mobility scaling factor over time (burn-in and analysis period)**

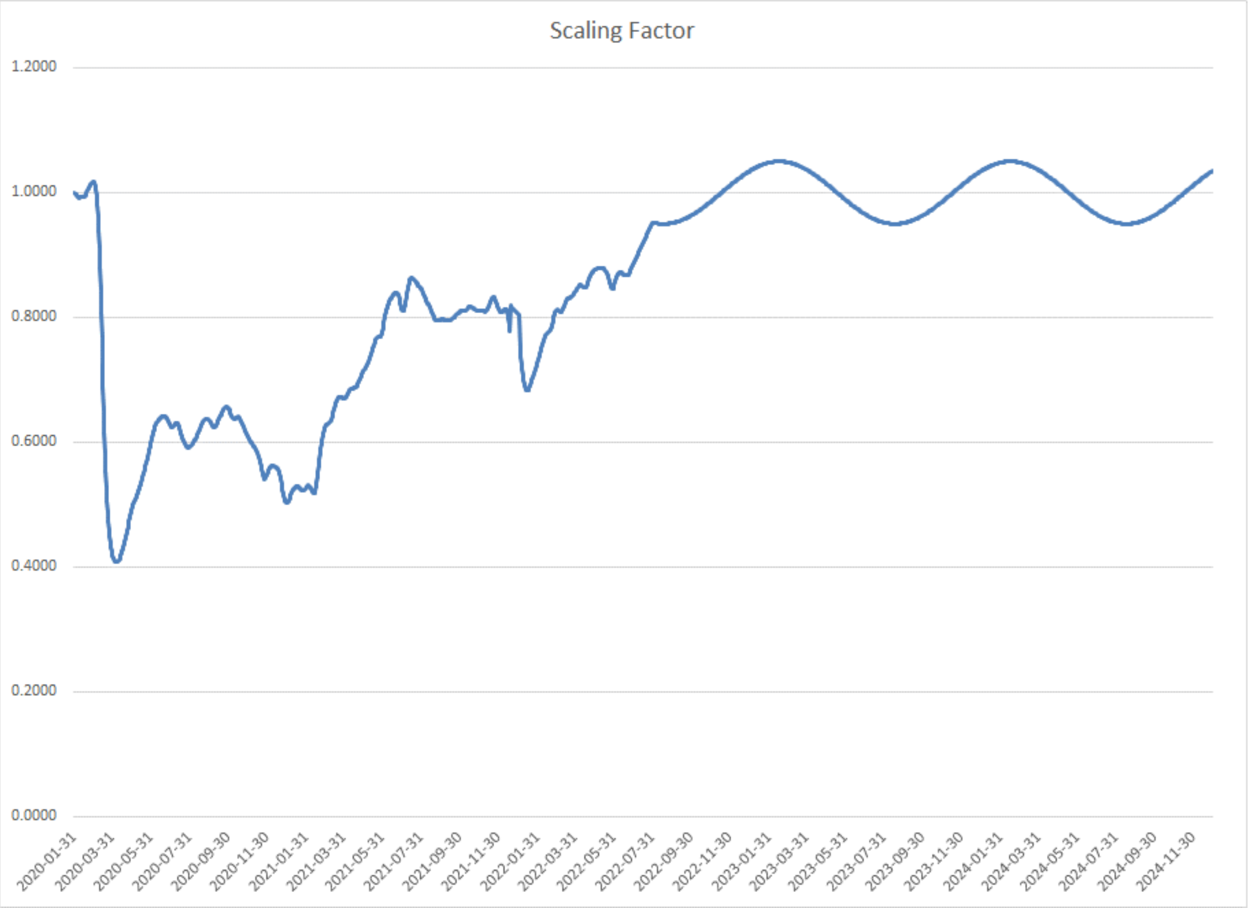

### 1.3. Reduction in Effective Contacts Due to Vaccination

#### 1.3.1. Vaccine Coverage

A proportion of the population moves into the primary series and booster model strata according to vaccine uptake data from the CDC, as illustrated in Figure 2 to Figure 5[7].

**Figure 2. Percent of the population who have completed their primary series, by age group**

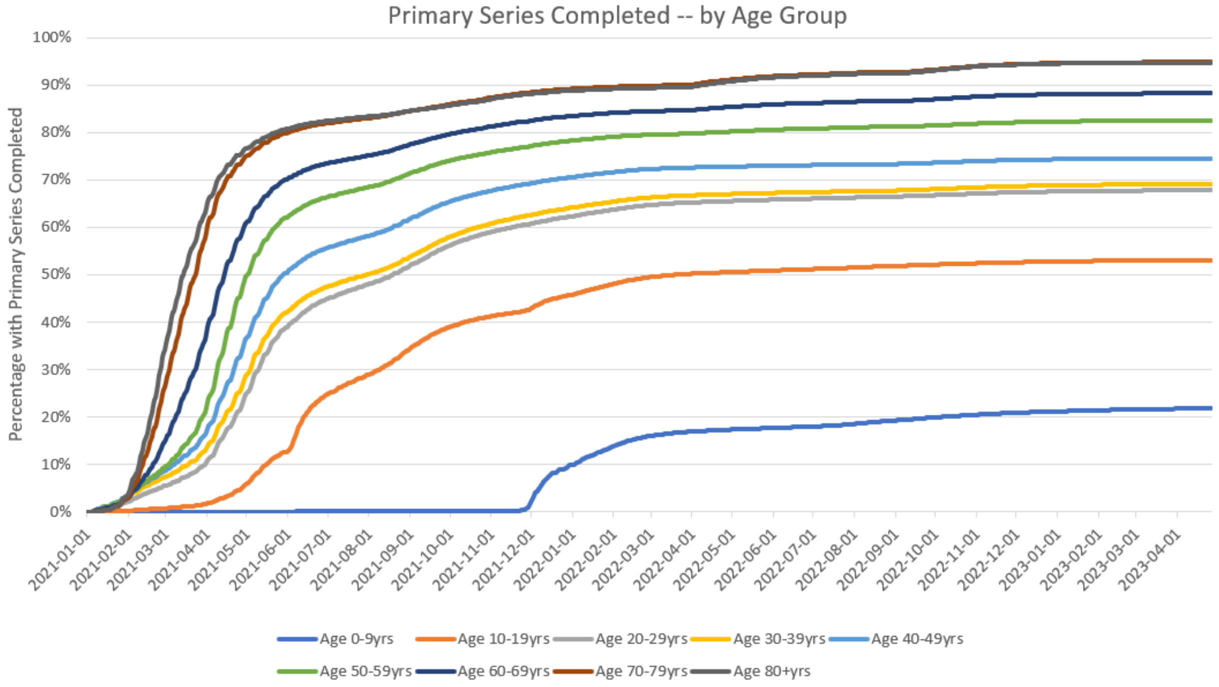

**Figure 3. Percent of the population who have received a booster, by age group**

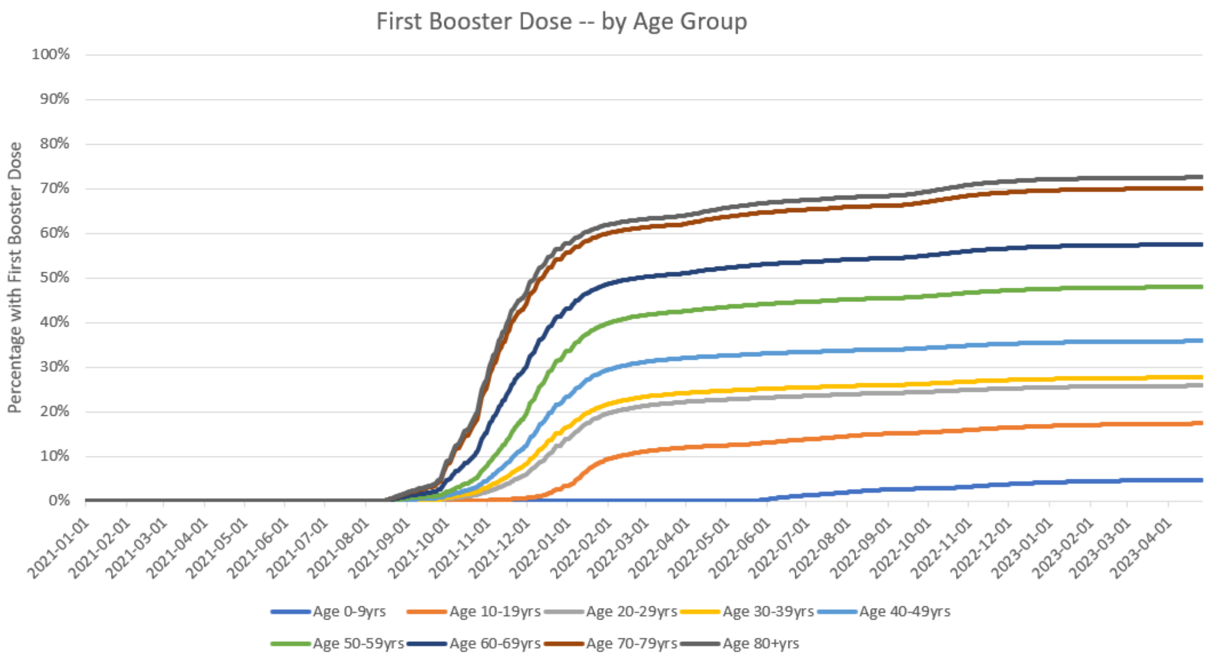

**Figure 4. Percent of the population who received two boosters, by age group**

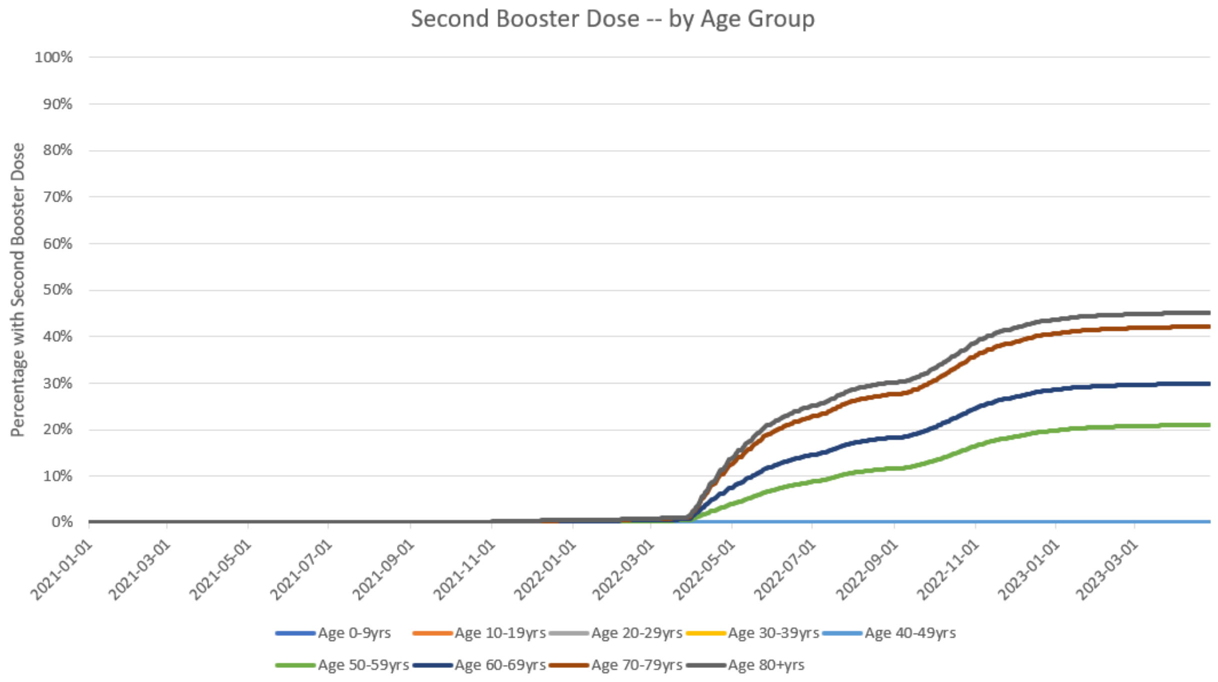

**Figure 5. Percent of the population who received a bivalent booster, by age group**

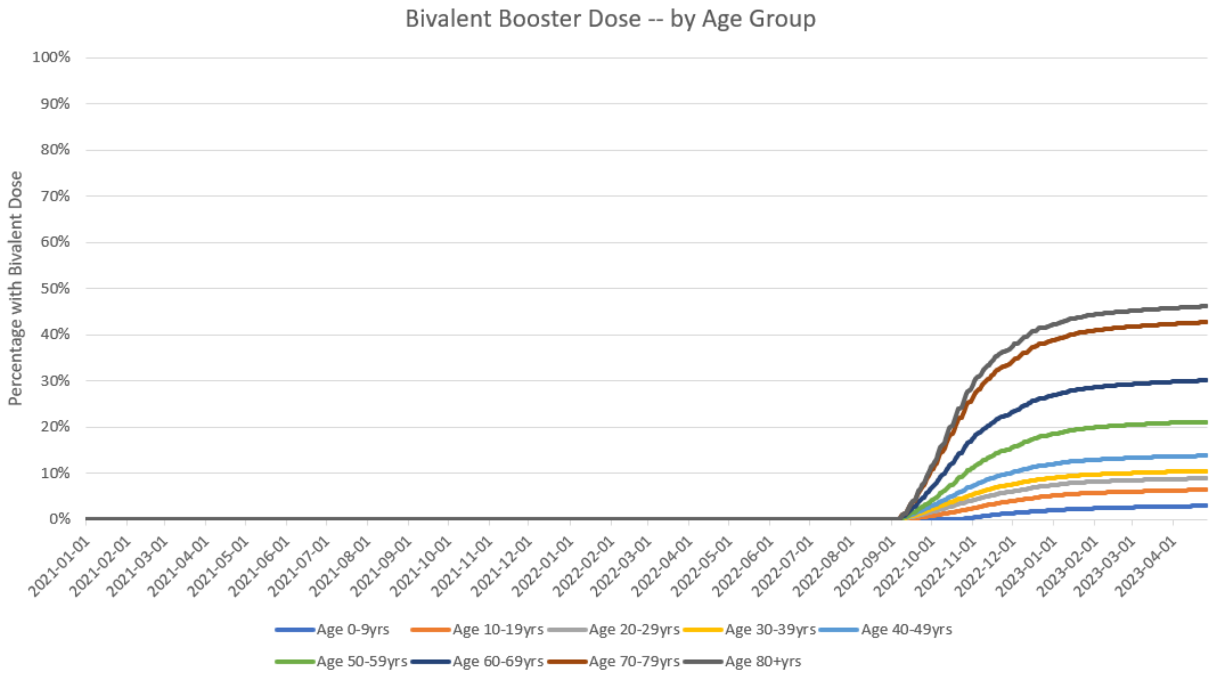

#### 1.3.2. Vaccine Effectiveness

To determine the residual VE of the target population, the initial VE and monthly waning for infection and severe disease for vaccines previously received by the population were required. These values differed by the vaccine, vaccination type (primary series or booster), and the circulating variant at the time of administration. Values used are provided below.

Delta

Primary series VE and monthly waning against the Delta variant for infection and severe disease for each vaccine were obtained from the Institute for Health Metrics and Evaluation (IHME) COVID-19 December 22, 2021 model update.[8] Booster VE and monthly waning for Spikevax (mRNA-1273, Moderna) and Comirnaty (BNT126b2, Pfizer-BioNTech) were obtained from Voko et al. (2021).[9] Because booster VE for Jcovden (Ad26.COV2.S, Janssen)) was not available, it was approximated by adjusting the primary series VE using the same ratio between primary series and booster observed with Comirnaty. Booster waning for infection and severe disease were assumed to be the same as primary series waning for infection and severe disease, respectively.

BA.1/BA.2

Primary series and booster VEs against BA.1/BA.2 for all vaccines for infection and severe disease were obtained from a meta-analysis by Pratama et al. (2022),[10] except for the Spikevax booster against severe disease, which was not available. Therefore, the ratio between booster infection VE to severe disease VE for Comirnaty was calculated and applied to the booster infection VE for Spikevax to approximate the booster severe disease VE for Spikevax. Waning against primary series VE was obtained from Pratama et al. (2022).[10] Booster waning against infection and severe disease were obtained from a meta-analysis by Higdon et al. (2022),[11] which examined the duration of booster VE over six months against the omicron variant.

BA.4/BA.5

Booster VE against infection and hospitalizations for the monovalent vaccines Spikevax and Comirnaty were approximated using an antibody titer model that was developed by Khoury et al. (2021)[12] and updated by Hogan et al. (2021).[13] Geometric mean titer (GMT) levels against ancestral, BA.1/2, and BA.4/5[14,15] induced by the vaccines were entered into the model, as well as known VE against infection for BA.1/2[10,16] to predict VE against infection and severe disease for BA.4/5.

GMT levels were not available for Jcovden. Therefore, the VE against infection and severe disease were approximated by using the relationship between the monovalent booster VEs against BA.1/BA.2 and BA.4/BA.5. Because the ratios in VEs against BA.1/BA.2 and BA.4/BA.5 differed for Spikevax and Comirnaty, an average of their ratios was used.

Booster VE against severe disease for the Spikevax bivalent vaccine was obtained from the Kaiser Permanente prospective cohort study by Tseng et al. (2023).[17] This study estimated the VE of the Spikevax bivalent booster (mRNA-1273.222) against severe disease caused by BA.4/BA.5. VE for infection was not estimated, and therefore was assumed to be the same as Spikevax monovalent against BA.1/2. To calculate the booster VE against severe disease for the Comirnaty bivalent vaccine (BNT126b2 (WT/OMI BA.4/BA.5)), data from a US-based, retrospective database analysis by Kopel et al. (2023) was used.[18] The relative vaccine effectiveness (rVE) against severe disease between the two mRNA bivalents was applied to the VE for Spikevax bivalent to approximate the VE against severe disease of Comirnaty bivalent. Booster VE against infections for both vaccines were assumed to be the same as monovalent booster against BA.1/BA.2.[10]

Booster waning for all vaccines against BA.4/BA.5 was assumed to be the same as monovalent vaccines against BA.1/BA.2 and obtained from Higdon et al. (2022).[11]

**Table 3. Starting vaccine effectiveness and monthly waning values for previously administered vaccinations.**

| | Primary series (%) | | | | Booster (%) | | | |
|---|---|---|---|---|---|---|---|---|
| | Infection | | Severe | | Infection | | Severe | |
| | VE | Waning | VE | Waning | VE | Waning | VE | Waning |
| **DELTA** | | | | | | | | |
| Spikevax | 91.0 | 3.4 | 97.0 | 1.0 | 93.3 | 3.4 | 95.7 | 1.0 |
| Comirnaty | 84.0 | 4.0 | 95.0 | 1.3 | 88.1 | 4.0 | 93.0 | 1.3 |
| Jcovden | 64.0 | 4.0 | 76.0 | 2.1 | 67.1 | 4.0 | 74.4 | 2.1 |
| **BA.1/BA.2** | | | | | | | | |
| Spikevax | 47.8 | 3.0 | 66.9 | 1.4 | 57.1 | 4.8 | 95.9 | 1.4 |
| Comirnaty | 52.2 | 3.4 | 80.2 | 1.4 | 53.0 | 4.8 | 89.1 | 1.4 |
| Jcovden | 26.4 | 5.3 | 31.0 | 3.5 | 48.7 | 4.8 | 81.7 | 1.4 |
| **BA.4/BA.5** | | | | | | | | |
| Spikevax | | | | | 32.3 | 4.8 | 62.6 | 1.4 |
| Comirnaty | | | | | 23.9 | 4.8 | 52.7 | 1.4 |
| Jcovden | | | | | 24.7 | 4.8 | 50.9 | 1.4 |
| Spikevax bivalent | | | | | 57.1 | 4.8 | 84.3 | 1.4 |
| Comirnaty bivalent | | | | | 53.0 | 4.8 | 82.6 | 1.4 |

VE: Vaccine effectiveness

US market shares[19] were applied to the VE and waning rates, by variant, to calculate a weighted VE value against each variant. These weighted values were entered into the SEIR model (Table 4).

**Table 4. Starting vaccine effectiveness and monthly waning values for previously administered vaccinations.**

| | Delta | | | | BA.1/BA.2 | | | | BA.4/BA.5 | | | |
|---|---|---|---|---|---|---|---|---|---|---|---|---|
| | Infection | | Severe | | Infection | | Severe | | Infection | | Severe | |
| | VE | Waning | VE | Waning | VE | Waning | VE | Waning | VE | Waning | VE | Waning |
| Primary series | 84.9 | 3.8 | 94.2 | 1.3 | 48.6 | 3.4 | 71.5 | 1.5 | | | | |
| First Booster | 90.0 | 3.7 | 93.9 | 1.2 | 54.7 | 4.8 | 91.9 | 1.4 | 27.5 | 4.8 | 56.9 | 1.4 |
| Second Booster | 90.3 | 3.7 | 94.1 | 1.2 | 54.7 | 4.8 | 91.9 | 1.4 | 27.4 | 4.8 | 56.8 | 1.4 |
| Bivalent Booster | | | | | | | | | 54.5 | 4.8 | 83.2 | 1.4 |

On the day that the Omicron BA.1 was emergent (December 13, 2021) and Omicron BA.4/BA.5 was emergent (June 6, 2022), it was assumed that protection from both vaccine-mediated and natural immunity dropped immediately. The decrease was calculated as the ratio of the vaccine effectiveness during each period.

### 1.4. Other Model Inputs

The average length of time spent by infected individuals exposed to the virus before they become infectious (e.g. latent period) was assumed to be 3 days.[20] The length of time spent by an individual in an infectious state (e.g. infectious period) was assumed to be 7 days.[20,21]

In the base case, the rate of natural immunity waning is set to be equal to the rate of waning of primary series VE against infection. Therefore, the monthly rate of waning is 3.8% for the pre-Omicron period and 3.4% for the Omicron and Omicron BA.4/5 periods. When vaccine-mediated immunity decreased on the days that the circulating variant changed (start of the Omicron period; start of the Omicron BA.4/5 period), a similar drop in the number of people in the Recovered compartment was also implemented.

### 1.5. Transmissibility: February 2020 to August 2023

During the first 30 days of the burn-in period, an initial 93,372infections used to seed or start the pandemic. This number is equal to the number of infections estimated by the Institute for Health Metrics and Evaluation (IHME) over the first 30 days.[22] Model calibration was then conducted to estimate the transmissibility parameter that reflects cases of COVID-19 experienced in the US from February 2020 through August 2023. The analytical choices for the calibration process were made considering the recommendations of Vanni and colleagues[23]. The transmissibility parameter was allowed to vary on a daily basis. For the calibration process, transmissibility parameters were manually varied. The calibration targets and the goodness of fit measures used, are described along with the results of the calibration below.

#### 1.5.1. Calibration Targets

As the collection of COVID-19 related infections has changed over time, two different outcomes of interest, or targets, were used for the calibration process.

For the first part of the pandemic, the target was the total number of infections in the population. This includes all asymptomatic and symptomatic infections, whether or not they have been reported. While data on the number of infections are collected by public health authorities, these figures reflect the number of reported infections and not the true number of infections. There are multiple factors that lead to under-testing and under-reporting of infections including patient and physician behavior as well as technical limitations to testing. Therefore, output from the modelling done by the IHME, which has been corrected for these biases and includes all infections (symptomatic and asymptomatic), was used as the calibration target. The IHME model was chosen because it was one of the models considered by the United States Centers for Disease Control over time in their ensemble model forecasts.[24] Prior to the pandemic, IHME had developed methods of collecting data globally, which they were able to do in real time during the pandemic in order to create projections for all countries across the globe. IHME has produced multiple publications on their COVID-19 model and the structural and input modifications over time.[25] Finally, the detailed outputs from their model are publicly available. The calibration target was the daily incidence of reported COVID-19 cases (symptomatic and asymptomatic), as reported by the IHME during the period of February 4, 2020 through March 31, 2022.[22] While IHME reported outputs after March 31, 2022, the overall reporting of infections became less reliable overall. Therefore, the endpoint used for this calibration changed after this date.

For the time period April 1, 2022 through March 31, 2023, the calibration target was the monthly number of hospitalizations, estimated from the hospitalization admission rates observed from April 2022 through March 2023 from the CDC's COVID-NET data.[26] The transmissibility parameters were changed manually to estimate the number of infections and the resultant number of hospitalizations. The overall monthly hospitalization rates per 100,000 population and the corresponding number of hospitalizations, multiplied by the population estimates, are presented in Table 5.

**Table 5. Target hospitalization rates and number of hospitalizations**

| Month | Rate per 100,000 population | Number of Hospitalizations |
|---|---|---|
| Apr 2022 | 15.0 | 50,391 |
| May 2022 | 31.2 | 104,670 |
| Jun 2022 | 33.5 | 112,445 |
| Jul 2022 | 41.9 | 140,616 |
| Aug 2022 | 39.1 | 131,497 |
| Sep 2022 | 28.3 | 95,120 |
| Oct 2022 | 27.8 | 93,296 |
| Nov 2022 | 31.9 | 107,166 |
| Dec 2022 | 43.4 | 145,751 |
| Jan 2023 | 34.3 | 115,276 |
| Feb 2023 | 24.1 | 80,962 |
| Mar 2023 | 19.4 | 65,077 |

#### 1.5.2. Goodness of Fit Measure

An initial model calibration was performed in a qualitative way by visually comparing the daily incidence of COVID cases predicted by the model and the values obtained from the IHME data for the first time period (February 4, 2020 through March 31, 2022). For the second time period of the calibration, April 2022 through March 2023, the monthly estimates of the number of hospitalizations from the model was to differ by no more than 10% from the target number of hospitalizations for each month.

#### 1.5.3. Results of the Calibration

The final transmissibility parameters for the calibration period for the base case scenario are summarized in Table 6. For the dates shown, the transmissibility parameter was manually varied and linear interpolation was used to estimate values for the days between these dates. The comparison plot of the daily number of incident cases of COVID predicted from the calibrated model and the corresponding estimates from the IHME is presented in Figure 6. The comparison of the monthly estimates of the number of hospitalizations is presented in Figure 7.

Table 6. Final transmissibility parameter inputs for the SEIR model

| Date | Days from start of analysis | Transmissibility parameter estimate |
|---|---|---|
| January 31, 2020 | 1 | 0.08 |
| February 14, 2020 | 15 | 0.08 |
| February 29, 2020 | 30 | 0.15 |
| March 15, 2020 | 45 | 0.15 |
| March 30, 2020 | 60 | 0.15 |
| April 14, 2020 | 75 | 0.15 |
| April 29, 2020 | 90 | 0.10 |
| May 14, 2020 | 105 | 0.10 |
| May 29, 2020 | 120 | 0.12 |
| June 13, 2020 | 135 | 0.12 |
| June 28, 2020 | 150 | 0.15 |
| July 13, 2020 | 165 | 0.12 |
| July 28, 2020 | 180 | 0.11 |
| August 12, 2020 | 195 | 0.10 |
| August 27, 2020 | 210 | 0.10 |
| September 11, 2020 | 225 | 0.11 |
| September 26, 2020 | 240 | 0.12 |
| October 11, 2020 | 255 | 0.14 |

| Date | Days from start of analysis | Transmissibility parameter estimate |
|---|---|---|
| October 26, 2020 | 270 | 0.16 |
| November 10, 2020 | 285 | 0.15 |
| November 25, 2020 | 300 | 0.15 |
| December 10, 2020 | 315 | 0.14 |
| December 25, 2020 | 330 | 0.16 |
| January 9, 2021 | 345 | 0.15 |
| January 24, 2021 | 360 | 0.12 |
| February 8, 2021 | 375 | 0.12 |
| February 23, 2021 | 390 | 0.14 |
| March 10, 2021 | 405 | 0.14 |
| March 25, 2021 | 420 | 0.16 |
| April 9, 2021 | 435 | 0.15 |
| April 24, 2021 | 450 | 0.14 |
| May 9, 2021 | 465 | 0.12 |
| May 24, 2021 | 480 | 0.13 |
| June 8, 2021 | 495 | 0.13 |
| June 23, 2021 | 510 | 0.18 |
| July 8, 2021 | 525 | 0.22 |
| July 23, 2021 | 540 | 0.22 |
| August 7, 2021 | 555 | 0.21 |
| August 22, 2021 | 570 | 0.19 |
| September 6, 2021 | 585 | 0.17 |
| September 21, 2021 | 600 | 0.16 |
| October 6, 2021 | 615 | 0.16 |
| October 21, 2021 | 630 | 0.20 |
| November 5, 2021 | 645 | 0.20 |
| November 20, 2021 | 660 | 0.33 |
| December 5, 2021 | 675 | 0.33 |
| December 20, 2021 | 690 | 0.31 |
| January 4, 2022 | 705 | 0.30 |
| January 19, 2022 | 720 | 0.30 |
| February 3, 2022 | 735 | 0.35 |
| February 18, 2022 | 750 | 0.40 |
| March 5, 2022 | 765 | 0.45 |
| March 20, 2022 | 780 | 0.55 |
| April 4, 2022 | 795 | 0.33 |
| April 19, 2022 | 810 | 0.30 |
| May 4, 2022 | 825 | 0.60 |
| May 19, 2022 | 840 | 0.88 |
| June 4, 2022 | 856 | 0.49 |
| June 7, 2022 | 859 | 0.14 |
| June 13, 2022 | 865 | 0.16 |
| June 18, 2022 | 870 | 0.16 |
| July 3, 2022 | 885 | 0.16 |
| July 18, 2022 | 900 | 0.15 |
| August 2, 2022 | 915 | 0.14 |
| August 17, 2022 | 930 | 0.14 |

| Date | Days from start of analysis | Transmissibility parameter estimate |
|---|---|---|
| September 1, 2022 | 945 | 0.14 |
| September 16, 2022 | 960 | 0.13 |
| October 1, 2022 | 975 | 0.14 |
| October 16, 2022 | 990 | 0.15 |
| October 31, 2022 | 1005 | 0.17 |
| November 15, 2022 | 1020 | 0.18 |
| November 23, 2022 | 1028 | 0.15 |
| November 30, 2022 | 1035 | 0.17 |
| December 16, 2022 | 1051 | 0.19 |
| December 31, 2022 | 1066 | 0.16 |
| January 16, 2023 | 1082 | 0.15 |
| January 31, 2023 | 1097 | 0.15 |
| February 14, 2023 | 1111 | 0.16 |
| February 28, 2023 | 1125 | 0.17 |
| March 31, 2023 | 1156 | 0.15 |
| April 30, 2023 | 1186 | 0.15 |
| May 31, 2023 | 1217 | 0.19 |

**Figure 6. Comparison of daily incidence of COVID cases (calibrated model estimates vs. IHME estimate)**

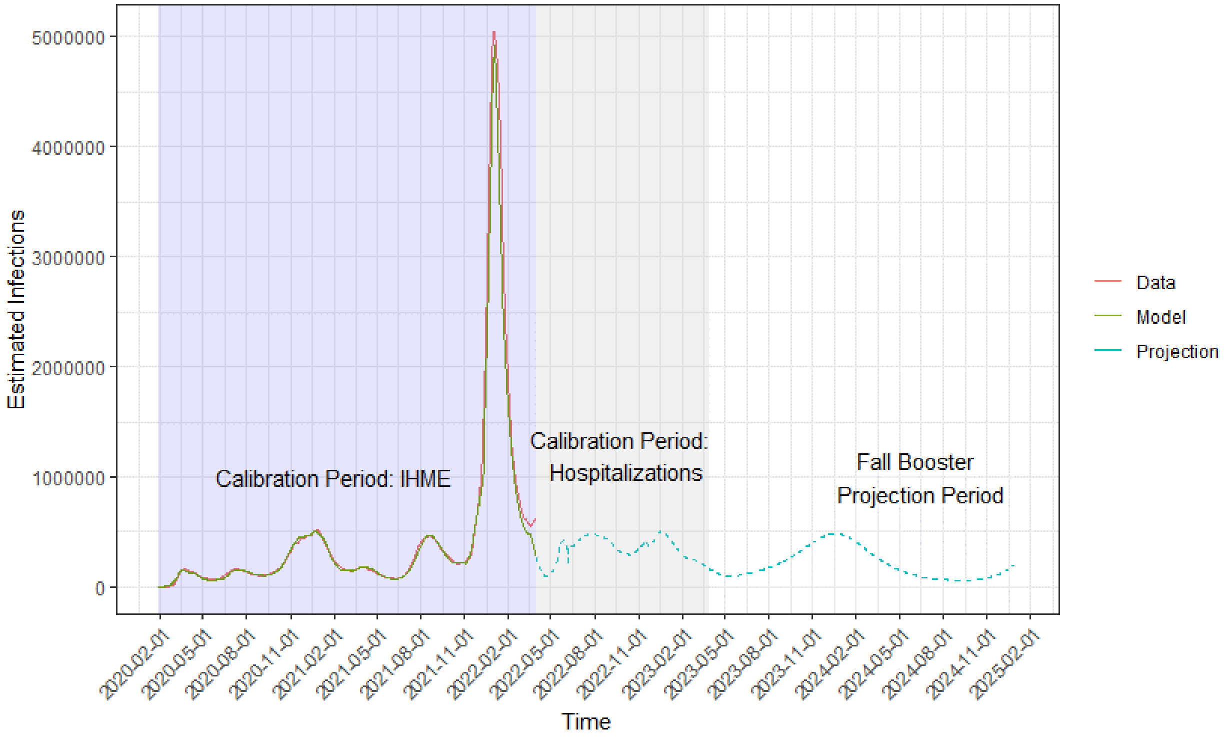

IHME: Institute for Health Metrics and Evaluation

**Figure 7. Comparison of monthly number of hospitalizations (calibrated model estimates vs. National Hospital Safety Network estimates)**

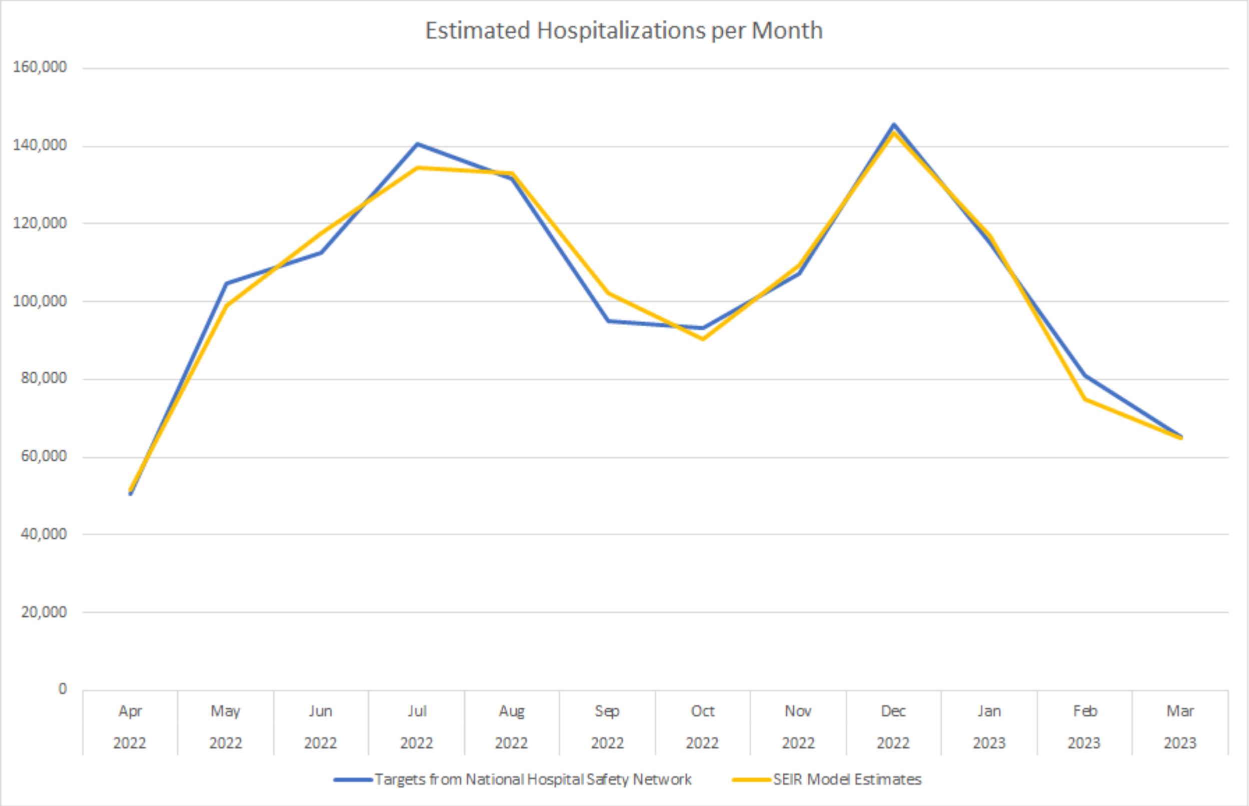

### 2. SEIR MODEL INPUTS: ANALYTIC TIME PERIOD (SEPTEMBER 2023 TO AUGUST 2024)

The inputs that were changed for the analytic time period are described below. Inputs not described were the same as during the burn-in period.

#### 2.1. Vaccine Coverage

Individuals under 18 years of age are modeled, but their vaccination status does not change after September 1, 2023, and they are not eligible to receive a fall booster as this analysis focuses on adults only.

Base case Fall 2023 vaccine uptake for those aged ≥18 years was based on influenza vaccine uptake for the 2022-2023 season and assumed to occur between September 2023-January 2024 (Figure 8). For the comparison of the Moderna strategy to the No Vaccine strategy, two scenario analyses were also included: one where the vaccine coverage is half of the base case values

(Figure 9), and one where coverage is lower than the base case, especially in the younger age group, as was observed for the bivalent booster (Figure 10).

**Figure 8. Fall 2023 vaccine coverage, base case**

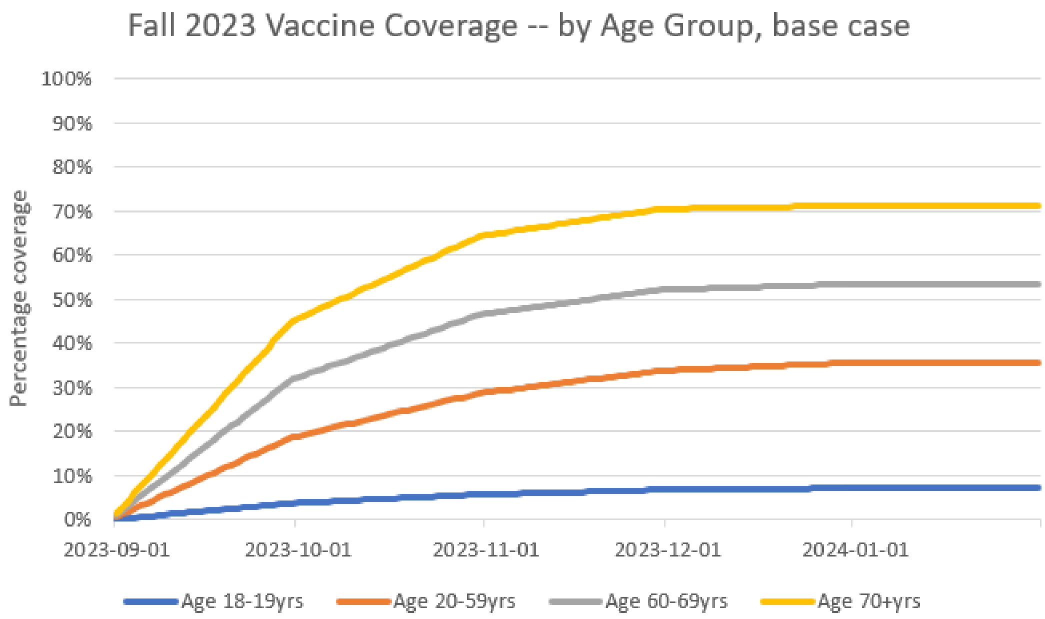

**Figure 9. Fall 2023 vaccine coverage, scenario analysis: half of base case values**

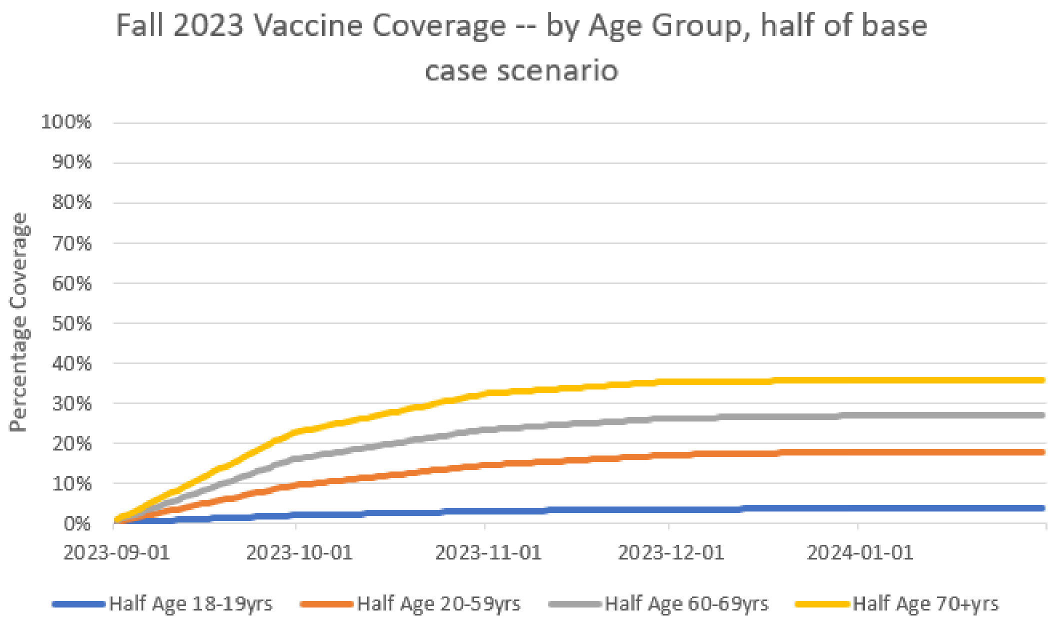

\

**Figure 10. Fall 2023 vaccine coverage, scenario analysis: uptake based on bivalent booster uptake pattern.**

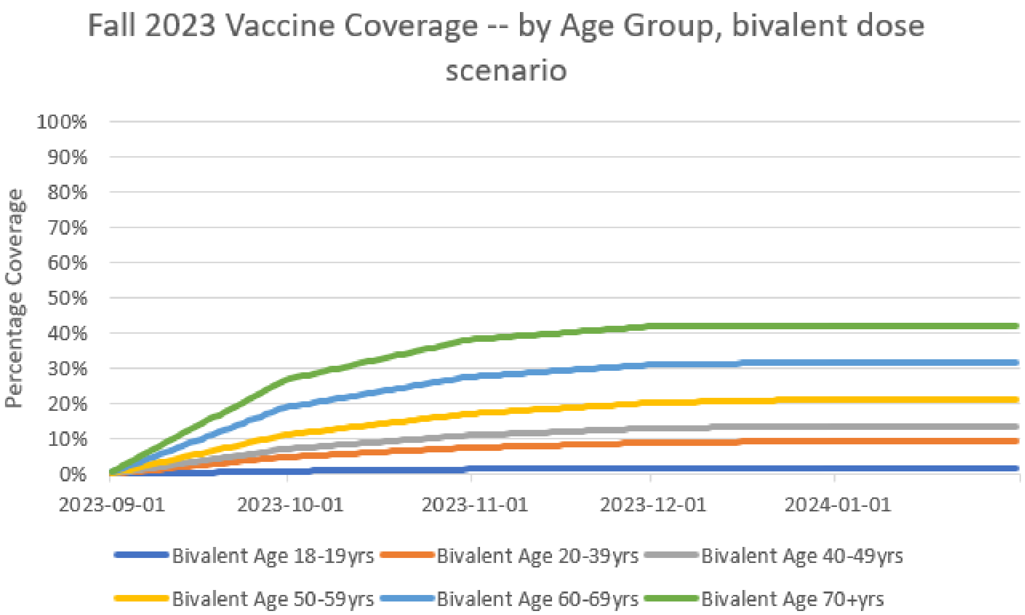

### 2.2. Vaccine Effectiveness

The Moderna updated COVID-19 mRNA Fall 2023 vaccine and the Pfizer updated COVID-19 mRNA Fall 2023 vaccine are assumed to be well-matched to the circulating variant at the time. Therefore, the VE of the Moderna bivalent against BA.4/BA.5 was assumed for the VE of the upcoming Moderna updated COVID-19 mRNA Fall 2023 vaccine. The rVEs estimated by Kopel et al. (2023) between the Moderna and Pfizer bivalent vaccines were used to estimate the VEs against severe disease and infection for the Pfizer updated COVID-19 mRNA Fall 2023 vaccine.[18] The rVE against hospitalization was used for severe disease. Because there were no data on infections, the rVE against outpatient visit was used as a proxy. Waning was assumed to be the same as monovalent vaccines against BA.1/BA.2. rVE values were available for the aged ≥65 years population and were therefore applied to the Moderna updated Fall 2023 vaccine VE values to estimate Pfizer updated Fall 2023 vaccine VE values for the scenario analysis.

**Table 7. Updated COVID-19 mRNA Fall 2023 vaccine initial vaccine effectiveness**

| | Infection (%) | Severe Disease (%) |
|---|---|---|
| Moderna updated COVID-19 mRNA Fall 2023 vaccine | 57.1 | 84.3 |
| Pfizer updated COVID-19 mRNA Fall 2023 vaccine | | |
| Base case: aged ≥18 years | 54.8 | 82.6 |
| Scenario analysis: aged ≥65 years | 52.0 | 81.9 |

VE: vaccine effectiveness

#### 2.3. Transmissibility: April 1, 2023 onwards

The values for the transmissibility parameter after September 1, 2023, were held constant at the final value in the calibration period (0.19) for the projections, assuming it represents the transmissibility of the variability of the currently circulating variant.

### 3. INFECTION CONSEQUENCES MODEL

Additional information on the inputs for the infection consequences model are provided in this section.

The table below includes the inputs for the societal perspective and key assumptions utilized in this scenario analysis, including time loss from work due to acute infection and hospitalization recovery. The proportions of patients with long COVID and severe long COVID are also presented, as severe long COVID patients are assumed to experience time loss.

**Table 8. Additional (non-healthcare related) inputs for the societal perspective scenario analysis**

| Model parameter | Value |
|---|---|
| Percentage in labor force[27] | |
| 18-29 years | 66.4% |
| 30-39 years | 78.1% |
| 40-49 years | 78.5% |
| 50-64 years | 66.5% |
| 65-74 years | 24.6% |
| 75-84 years | 8.3% |
| ≥85 years | 8.3% |
| Proportion with long COVID[28] | |
| 18-29 years | 26.9% |
| 30-39 years | 31.7% |
| 40-49 years | 32.6% |
| 50-64 years | 32.3% |
| 65-74 years | 28.0% |
| 75-84 years | 22.7% |
| ≥85 years | 18.5% |

| | |
|---|---|
| Proportion with severe long COVID (of those with long COVID)[29] | 6.9% |
| Daily wage[30] | $261 |
| Days lost[a]: | |
| Vaccination[31,32b] | 0.04 |
| Symptomatic infection, not hospitalized[33c] | 3.57 |
| Symptomatic infection, hospitalized[34d]: | |
| No ICU or ventilator | 4.36 |
| ICU only | 6.86 |
| ICU with ventilator | 12.07 |
| Hospitalization recovery[35d] | 33.43 |
| Severe long COVID[29] | 24.66 |

[a] Days lost may not fall only on work days; estimates were adjusted by 5/7 to account for this
[b]Assumes 95%[16] of patients receive vaccine in a pharmacy setting (0.2 hours of time lost[32]) and 5% receive vaccine in a physician's office setting (2 hours of time lost[32]).
[c]Applies to all non-hospitalized patients, including those who did not seek outpatient care.
[d]Hospitalized patients incur 3.57 days of time loss (from symptomatic infection, not hospitalized) to capture time loss due to symptoms prior to hospitalization, and an additional 33.43 days of time loss which represents time loss up to 60 days post-discharge.

**Table 9. The inputs for the vaccine-related adverse events: base case values**

| Model Parameter | Value |
|---|---|
| **Rates** | |
| Grade 3 local[36a,b] | 4.90% |
| Grade 3 systemic[36a,b] | 7.61% |
| Anaphylaxis[37c] | 0.0005% |
| Vaccine-induced myocarditis/pericarditis[38d] | 0.0018% |
| **Costs** | |
| Grade 3 local[39,40] | $7.59 |
| Grade 3 systemic[39] | $11.49 |
| Anaphylaxis[39,41,42,43,44] | $3,413 |
| Vaccine-induced myocarditis/pericarditis[39,45] | $3,584 |
| **QALYs Lost[32]** | |
| Grade 3 local | 0.0003 |
| Grade 3 systemic | 0.0011 |
| Anaphylaxis | 0.0019 |
| Vaccine-induced myocarditis/pericarditis | 0.0019 |

[a]Rates are from the AEs for monovalent boosters and assumed to apply to future vaccine versions. Grade 4 adverse events were not included (there were no grade 4 adverse events [local or systemic] reported in the clinical trial.

[b]Adverse event rates were assumed to be the same between Moderna and Pfizer.

[c]Vaccine-specific rates had overlapping 95% CI. Therefore, Moderna assumed to be the same as Pfizer.

[d]Vaccine-specific rates had overlapping 95% CI. Therefore, reported pooled data for Moderna and Pfizer used. Sex-specific pooled rates weighted by proportion of male and female US population aged 18-39. Risk applies to those ages 18-39 years only.

### 4. ADDITIONAL RESULTS TABLES AND FIGURES

#### Table 10. Disaggregated economic results

| Parameter | No Fall 2023 Vaccine (in millions) | Moderna updated Fall 2023 vaccine (in millions) | Pfizer-BioNTech updated Fall 2023 vaccine (in millions) | Δ Moderna Strategy – No Fall 2023 Vaccine Strategy (in millions) | Δ Moderna Strategy – Pfizer-BioNTech Strategy (in millions) |
|---|---|---|---|---|---|
| **Age ≥18 years** | | | | | |
| Vaccinations | $0 | $16,646 | $16,646 | $16,646 | $0 |
| Adverse events | $0 | $142 | $142 | $142 | $0 |
| Short-term infection | $39,731 | $30,666 | $31,381 | -$9,066 | -$716 |
| Post-infection | $12,536 | $10,660 | $10,872 | -$1,876 | -$211 |
| Infection related myocarditis | $987 | $862 | $877 | -$125 | -$15 |
| Productivity loss | $38,792 | $34,607 | $35,242 | -$4,186 | -$636 |
| **Total (healthcare perspective)** | **$53,255** | **$58,976** | **$59,918** | **$5,722** | **-$942** |
| **Total (societal perspective)** | **$92,047** | **$93,583** | **$95,160** | **$1,536** | **-$1,577** |
| **Age ≥65 years** | | | | | |
| Vaccinations | $0 | $5,962 | $5,962 | $5,962 | $0 |
| Adverse events | $0 | $50 | $50 | $50 | $0 |
| Short-term infection | $39,731 | $35,206 | $35,783 | -$4,525 | -$576 |
| Post-infection | $12,536 | $11,728 | $11,859 | -$809 | -$132 |
| Infection related myocarditis | $987 | $932 | $941 | -$55 | -$9 |
| Productivity loss | $38,792 | $37,124 | $37,355 | -$1,669 | -$231 |
| **Total (healthcare perspective)** | **$53,255** | **$53,879** | **$54,596** | **$624** | **-$717** |
| **Total (societal perspective)** | **$92,047** | **$91,003** | **$91,951** | **-$1,044** | **-$949** |

Table 10 shows the disaggregated economic results from the base case analysis. With the Moderna updated COVID-19 mRNA Fall 2023 vaccine, the model predicted that there will be $10,924 million in health care treatment costs prevented (or $98.33 treatment cost saved per vaccination) compared to No Fall 2023 vaccine. A sensitivity analyses was conducted with the unit price of the Moderna updated COVID-19 mRNA Fall 2023 vaccine unit cost. If the unit cost of the vaccine is increased from $129.50 to $411.44, the incremental cost-effectiveness ratio (ICER) for the Moderna vaccine compared to no vaccine will be $50,000 per quality-adjusted life-years (QALYs) gained. If the unit cost is increased to $744.87 or $1078.31 the cost per QALY gained will be $100,000 and $150,000 respectively. The results of additional sensitivity analyses are shown in Table 11 below.

**Table 11. Deterministic sensitivity analysis results (Moderna Updated Fall 2023 Vaccine relative to No Fall 2023 Vaccine)**

| Model Parameter | Variation | ICER (Cost/QALY Gained) | | Change from Base Case | | Change from Base Case (%) | |
|---|---|---|---|---|---|---|---|
| | | Low Value | High Value | Low Value | High Value | Low Value | High Value |
| Percentage infected with symptoms | 95% CI | $10,404 | $5,561 | $2,681 | -$2,161 | 35% | -28% |
| Societal perspective | Societal | $2,073 | | -$5,650 | | -73% | |
| Infection induced myocarditis rates | 95% CI | $7,753 | $7,684 | $31 | -$38 | 0.4% | -0.5% |
| Infection induced myocarditis cost | ±25% | $7,765 | $7,680 | $42 | -$42 | 0.5% | -0.5% |
| Infection induced myocarditis QALYs lost | ±25% | $7,723 | $7,723 | $0.04 | -$0.04 | 0.001% | -0.001% |
| Proportion seeking outpatient care | ±25% | $8,215 | $7,230 | $493 | -$493 | 6.4% | -6.4% |
| Total cost of outpatient care (per patient seeking) | ±25% | $8,215 | $7,230 | $493 | -$493 | 6.4% | -6.4% |
| Short-term infection period QALYS lost | ±25% | $7,789 | $7,657 | $67 | -$66 | 0.9% | -0.9% |
| Hospitalization rates (unvaccinated)* | 95% CI | $12,148 | $3,083 | $4,425 | -$4,640 | 57.3% | -60.1% |
| Percentage in ICU and ICU with Ventilator | 95% CI | $7,838 | $7,610 | $116 | -$113 | 1.5% | -1.5% |
| Hospitalization cost per stay | ±25% | $10,176 | $5,269 | $2,454 | -$2,454 | 31.8% | -31.8% |
| Hospitalization recovery cost | ±25% | $7,835 | $7,610 | $113 | -$113 | 1.5% | -1.5% |
| In-hospital mortality rates | 95% CI | $7,841 | $7,608 | $118 | -$115 | 1.5% | -1.5% |
| Percentage with hospital readmission | 95% CI | $7,752 | $7,693 | $30 | -$30 | 0.4% | -0.4% |
| Post-discharge mortality | ±25% | $8,334 | $7,195 | $611 | -$527 | 7.9% | -6.8% |
| Post-infection QALYS lost | 95% CI or ±25% | $8,473 | $7,078 | $750 | -$645 | 9.7% | -8.4% |
| Post-infection costs | ±25% | $8,356 | $7,090 | $633 | -$633 | 8.2% | -8.2% |
| Incidence | Double the waning rate for natural immunity during Omicron period | $2,400 | | -$5,323 | | -69% | |

| | | | | |
|---|---|---|---|---|
| Incidence | Half the waning rate for natural immunity during Omicron period | $7,642 | -$80 | -1.0% |
| Incidence | Emergence of XBB variant with immune escape in January 2023 | $38,749 | $31,026 | 402% |
| Vaccine coverage | Uptake half of base-case | $6,192 | -$1,531 | -20% |
| Vaccine coverage | Bivalent dose coverage | $3,943 | -$3,780 | -49% |
| Moderna Fall 2023 COVID-19 vaccine effectiveness | Decreased Moderna Fall 2023 vaccine initial VE (Lower 95% CI) against infection | $27,082 | $19,360 | 251% |
| Moderna Fall 2023 COVID-19 vaccine effectiveness | Increased Moderna Fall 2023 vaccine initial VE (Upper 95% CI) against infection | Fall 2023 booster dominates No Fall 2023 booster | | |
| Moderna Fall 2023 COVID-19 vaccine effectiveness | Decreased Moderna Fall 2023 vaccine initial VE (Lower 95% CI) against hospitalization | $9,193 | $1,471 | 19% |
| Moderna Fall 2023 COVID-19 vaccine effectiveness | Increased Moderna Fall 2023 vaccine initial VE (Upper 95% CI) against hospitalization | $6,672 | $1,050 | 14% |
| Moderna Fall 2023 COVID-19 vaccine effectiveness | Moderna vaccine initial VE (Lower 95% CI) against both infection and hospitalization | $33,196 | $25,474 | 330% |
| Moderna Fall 2023 COVID-19 vaccine effectiveness | Moderna vaccine initial VE (Upper 95% CI) against both infection and hospitalization | Fall 2023 booster dominates No Fall 2023 booster | | |
| Moderna Fall 2023 COVID-19 vaccine effectiveness | Decreased Fall 2023 vaccine waning (Outcome: Infection) | $1,213 | -$6,510 | -84% |
| Moderna Fall 2023 COVID-19 vaccine effectiveness | Increased Fall 2023 vaccine waning (Outcome: Infection) | $22,198 | $14,476 | 187% |
| Moderna Fall 2023 COVID-19 vaccine effectiveness | Decreased Fall 2023 vaccine waning | $6,369 | $1,354 | -18% |

| | | | | |
|---|---|---|---|---|
| | (outcome: hospitalization) | | | |
| Moderna Fall 2023 COVID-19 vaccine effectiveness | Increased Fall 2023 vaccine waning (outcome: hospitalization) | $9,697 | $1,974 | 26% |
| Moderna Fall 2023 COVID-19 vaccine effectiveness | Decreased Fall 2023 vaccine waning | $622 | -$7,101 | -92% |
| Moderna Fall 2023 COVID-19 vaccine effectiveness | Increased Fall 2023 vaccine waning | $28,739 | $21,016 | 272% |
| Moderna Fall 2023 COVID-19 vaccine effectiveness | Emergence of new variant with immune escape in November 2023 | $11,723 | $4,000 | 52% |
| Moderna Fall 2023 COVID-19 vaccine effectiveness | Emergence of new variant with immune escape in January 2024 | No Fall 2023 Booster dominates Fall 2023 Booster | | |
| Moderna Fall 2023 COVID-19 vaccine effectiveness | Emergence of new variant with immune escape in March 2024 | $54,448 | $46,726 | 605% |
| Target population | Ages 65+ only | $1,823 | -$5,900 | -76% |

*Reflects uncertainty in both the Omicron vs. Delta relative risk used to adjust the hospitalization rates for Omicron and the hospitalization rates

**Figure 11. Probabilistic sensitivity analyses: Cost-effectiveness acceptability curves.**

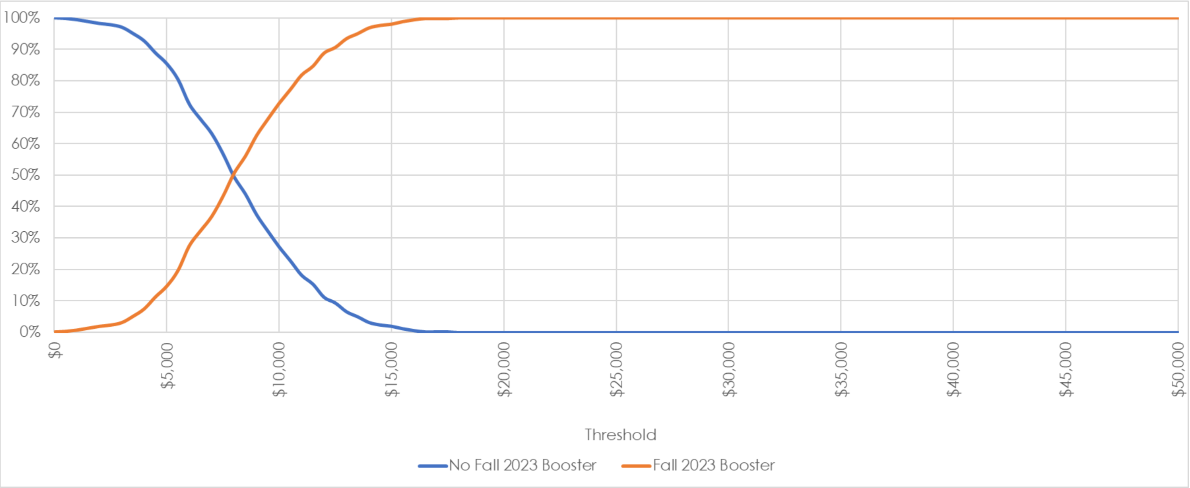

### 5. EQUATIONS ASSOCIATED WITH THE SEIR MODEL

#### 5.1. Differential Equations

The following equations describe the movement of people in the population through the SEIR compartments and the vaccination strata.

**Unvaccinated cohort**

$$S^X_{t+1,j} = S^X_{t,j} - \lambda^X_{t,j} S^X_{t,j} - \left(p^{X,S}_{t,j}\right)\mu_{t,j} + \omega^X_{t,j} R^X_{t,j} - \epsilon_{t,j}$$

$$E^X_{t+1,j} = E^X_{t,j} + \lambda^X_{t,j} S^X_{t,j} - \frac{1}{\tau_E} E^X_{t,j}$$

$$I^X_{t+1,j} = I^X_{t,j} + \frac{1}{\tau_E} E^X_{t,j} - \frac{1}{\tau_I} I^X_{t,j} + \epsilon_{t,j}$$

$$R^X_{t+1,j} = R^X_{t,j} - \left(p^{X,R}_{t,j}\right)\mu_{t,j} + \frac{1}{\tau_I} I^X_{t,j} - \omega^X_{t,j} R^X_{t,j}$$

**Vaccinated cohort**

$$S^V_{t+1,j} = S^V_{t,j} - \lambda^V_{t,j} S^V_{t,j} + \left(p^{X,S}_{t,j}\right)\mu_{t,j} - \left(p^{V,S}_{t,j}\right)\nu_{t,j} - \left(p^{V,S}_{t,j}\right) p^{V,B3}_{t,j} \nu^3_{t,j} - \left(p^{V,S}_{t,j}\right) p^{V,B4}_{t,j} \nu^4_{t,j} + \omega^V_{t,j} R^V_{t,j}$$

$$E^V_{t+1,j} = E^V_{t,j} + \lambda^V_{t,j} S^V_{t,j} - \frac{1}{\tau_E} E^V_{t,j}$$

$$I^V_{t+1,j} = I^V_{t,j} + \frac{1}{\tau_E} E^V_{t,j} - \frac{1}{\tau_I} I^V_{t,j}$$

$$R^V_{t+1,j} = R^V_{t,j} + \left(p^{X,R}_{t,j}\right)\mu_{t,j} - \left(p^{V,R}_{t,j}\right)\nu_{t,j} - \left(p^{V,R}_{t,j}\right) p^{V,B3}_{t,j} \nu^3_{t,j} - \left(p^{V,R}_{t,j}\right) p^{V,B4}_{t,j} \nu^4_{t,j} + \frac{1}{\tau_I} I^V_{t,j} - \omega^V_{t,j} R^V_{t,j}$$

**First Booster cohort**

$$S^B_{t+1,j} = S^B_{t,j} - \lambda^B_{t,j} S^B_{t,j} + \left(p^{V,S}_{t,j}\right)\nu_{t,j} - \left(p^{B,S}_{t,j}\right)\nu^2_{t,j} - \left(p^{B,S}_{t,j}\right)\left(p^{B,B3}_{t,j}\right)\nu^3_{t,j} - \left(p^{B,S}_{t,j}\right)\left(p^{B,B4}_{t,j}\right)\nu^4_{t,j} + \omega^B_{t,j} R^B_{t,j}$$

$$E^B_{t+1,j} = E^B_{t,j} + \lambda^B_{t,j} S^B_{t,j} - \frac{1}{\tau_E} E^B_{t,j}$$

| |
|---|
| $$I_{t+1,j}^{B} = I_{t,j}^{B} + \frac{1}{\tau_E} E_{t,j}^{B} - \frac{1}{\tau_I} I_{t,j}^{B}$$ $$R_{t+1,j}^{B} = R_{t,j}^{B} + (p_{t,j}^{V,R})\nu_{t,j} - (p_{t,j}^{B,R})\nu_{t,j}^{2} - (p_{t,j}^{B,R})(p_{t,j}^{B,B3})\nu_{t,j}^{3} - (p_{t,j}^{B,R})(p_{t,j}^{B,B4})\nu_{t,j}^{4} + \frac{1}{\tau_I} I_{t,j}^{B} - \omega_{t,j}^{B} R_{t,j}^{B}$$ |
| **Second Booster cohort** |
| $$S_{t+1,j}^{B2} = S_{t,j}^{B2} - \lambda_{t,j}^{B2} S_{t,j}^{B2} + (p_{t,j}^{B,S})\nu_{t,j}^{2} - (p_{t,j}^{B2,S})(p_{t,j}^{B2,B3})\nu_{t,j}^{3} - (p_{t,j}^{B2,S})(p_{t,j}^{B2,B4})\nu_{t,j}^{4} + \omega_{t,j}^{B2} R_{t,j}^{B2}$$ $$E_{t+1,j}^{B2} = E_{t,j}^{B2} + \lambda_{t,j}^{B2} S_{t,j}^{B2} - \frac{1}{\tau_E} E_{t,j}^{B2}$$ $$I_{t+1,j}^{B2} = I_{t,j}^{B2} + \frac{1}{\tau_E} E_{t,j}^{B2} - \frac{1}{\tau_I} I_{t,j}^{B2}$$ $$R_{t+1,j}^{B2} = R_{t,j}^{B2} + (p_{t,j}^{B,R})\nu_{t,j}^{2} - (p_{t,j}^{B2,R})(p_{t,j}^{B2,B3})\nu_{t,j}^{3} - (p_{t,j}^{B2,R})(p_{t,j}^{B2,B4})\nu_{t,j}^{4} + \frac{1}{\tau_I} I_{t,j}^{B2} - \omega_{t,j}^{B2} R_{t,j}^{B2}$$ |
| **Third Booster cohort** |
| $$S_{t+1,j}^{B3} = S_{t,j}^{B3} - \lambda_{t,j}^{B3} S_{t,j}^{B3} + \left((p_{t,j}^{V,S})p_{t,j}^{V,B3} + (p_{t,j}^{B,S})p_{t,j}^{B,B3} + (p_{t,j}^{B2,S})p_{t,j}^{B2,B3}\right)\nu_{t,j}^{3} - (p_{t,j}^{B3,S})(p_{t,j}^{B3,B4})\nu_{t,j}^{4} + \omega_{t,j}^{B3} R_{t,j}^{B3}$$ $$E_{t+1,j}^{B3} = E_{t,j}^{B3} + \lambda_{t,j}^{B3} S_{t,j}^{B3} - \frac{1}{\tau_E} E_{t,j}^{B3}$$ $$I_{t+1,j}^{B3} = I_{t,j}^{B3} + \frac{1}{\tau_E} E_{t,j}^{B3} - \frac{1}{\tau_I} I_{t,j}^{B3}$$ $$R_{t+1,j}^{B3} = R_{t,j}^{B3} + \left((p_{t,j}^{V,R})p_{t,j}^{V,B3} + (p_{t,j}^{B,R})p_{t,j}^{B,B3} + (p_{t,j}^{B2,R})p_{t,j}^{B2,B3}\right)\nu_{t,j}^{3} - (p_{t,j}^{B3,R})(p_{t,j}^{B3,B4})\nu_{t,j}^{4} + \frac{1}{\tau_I} I_{t,j}^{B3} - \omega_{t,j}^{B3} R_{t,j}^{B3}$$ |
| **Fourth Boosted cohort** |
| $$S_{t+1,j}^{B4} = S_{t,j}^{B4} - \lambda_{t,j}^{B4} S_{t,j}^{B4} + \left((p_{t,j}^{V,S})p_{t,j}^{V,B4} + (p_{t,j}^{B,S})p_{t,j}^{B,B4} + (p_{t,j}^{B2,S})p_{t,j}^{B2,B4} + (p_{t,j}^{B3,S})p_{t,j}^{B3,B4}\right)\nu_{t,j}^{4} + \omega_{t,j}^{B4} R_{t,j}^{B4}$$ $$E_{t+1,j}^{B4} = E_{t,j}^{B4} + \lambda_{t,j}^{B4} S_{t,j}^{B4} - \frac{1}{\tau_E} E_{t,j}^{B4}$$ |

$$I_{t+1,j}^{B4} = I_{t,j}^{B4} + \frac{1}{\tau_E} E_{t,j}^{B4} - \frac{1}{\tau_I} I_{t,j}^{B4}$$

$$R_{t+1,j}^{B4} = R_{t,j}^{B4} + \left( \left( p_{t,j}^{V,R} \right) p_{t,j}^{V,B4} + \left( p_{t,j}^{B,R} \right) p_{t,j}^{B,B4} + \left( p_{t,j}^{B2,R} \right) p_{t,j}^{B2,B4} + \left( p_{t,j}^{B3,R} \right) p_{t,j}^{B3,B4} \right) \nu_{t,j}^{4} + \frac{1}{\tau_I} I_{t,j}^{B4} - \omega_{t,j}^{B4} R_{t,j}^{B4}$$

**For t=1,**

$S_{1,j}^{X}$ = Initial number of susceptible individuals

$I_{1,j}^{X}$= Initial number of infectious individuals = $\epsilon_{1,j}$

$E_{1,j}^{X} = R_{1,j}^{X} = 0$

$S_{1,j}^{V} = E_{1,j}^{V} = I_{1,j}^{V} = R_{1,j}^{V} = 0$

$S_{1,j}^{B} = E_{1,j}^{B} = I_{1,j}^{B} = R_{1,j}^{B} = 0$

$S_{1,j}^{B2} = E_{1,j}^{B2} = I_{1,j}^{B2} = R_{1,j}^{B2} = 0$

$S_{1,j}^{B3} = E_{1,j}^{B3} = I_{1,j}^{B3} = R_{1,j}^{B3} = 0$

$S_{1,j}^{B4} = E_{1,j}^{B4} = I_{1,j}^{B4} = R_{1,j}^{B4} = 0$

Notes, superscripts:

*X*, *V*, *B*, *B2*, *B3*, and *B4* represent the unvaccinated, vaccinated, first booster, second booster, third booster (bivalent), and fourth booster (fall 2023 bivalent) cohorts, respectively.

Notes, subscripts:

*t* = time (i.e., day of analysis)

*j* = age group (number 1 to 9)

**Definitions**

$S_j^X, S_j^V, S_j^B, S_j^{B2}, S_j^{B3}, S_j^{B4}$ represent the proportion of susceptibles in age group *j* in cohort *X*, *V*, *B*, *B2*, *B3*, or *B4*.

The compartments with superscript *X*, *V*, *B, B1, B2, B3, and B4* represent the unvaccinated, vaccinated, and boosted cohorts, respectively.

| |
|---|
| $E_j^Z$ represent exposed, but not yet infectious, individuals in age group *j* in cohort *Z* (*X*, *V*, *B, B2*, *B3*, or *B4*).<br><br>$I_j^Z$ represent infectious individuals in age group *j* in cohort *Z* (*X*, *V*, *B, B2*, *B3*, or *B4*).<br><br>$R_j^Z$ represent immune individuals in age group *j* in cohort *Z* (*X*, *V*, *B, B2*, *B3*, or *B4*). |
| $\lambda_{t,j}^*$ is the age-group specific force of infection (see section below)<br><br>$\frac{1}{\tau_E}$ is the rate of loss of latency<br><br>$\frac{1}{\tau_I}$ is the rate of loss of infectiousness<br><br>$\mu_{t,j}$ is the proportion receiving a (primary series) vaccination on day *t* in age group *j*<br><br>$\nu_{t,j}$ is the proportion receiving a booster on day *t* in age group *j*<br><br>$\nu_{t,j}^2$ is the proportion receiving a second booster on day *t* in age group *j*<br><br>$\nu_{t,j}^3$ is the proportion receiving a third booster on day *t* in age group *j*<br><br>$\nu_{t,j}^4$ is the proportion receiving a fourth booster on day *t* in age group *j*<br><br>$p_{t,j}^{V,B3}$ is the proportion receiving a third booster from cohort *V* on day *t* in age group *j*<br><br>$p_{t,j}^{B,B3}$ is the proportion receiving a third booster from cohort *B* on day *t* in age group *j*<br><br>$p_{t,j}^{B2,B3}$ is the proportion receiving a third booster from cohort *B2* on day *t* in age group *j*<br><br>$p_{t,j}^{V,B4}$ is the proportion receiving a fourth booster from cohort *V* on day *t* in age group *j*<br><br>$p_{t,j}^{B,B4}$ is the proportion receiving a fourth booster from cohort *B* on day *t* in age group *j*<br><br>$p_{t,j}^{B2,B4}$ is the proportion receiving a fourth booster from cohort *B2* on day *t* in age group *j*<br><br>$p_{t,j}^{B3,B4}$ is the proportion receiving a fourth booster from cohort *B3* on day *t* in age group *j*<br><br>Note: $p_{t,j}^{V,B3} + p_{t,j}^{B,B3} + p_{t,j}^{B2,B3} = 1$<br><br>Note: $p_{t,j}^{V,B4} + p_{t,j}^{B,B4} + p_{t,j}^{B2,B4} + p_{t,j}^{B3,B4} = 1$<br><br>$p_{t-1,j}^{Z,S}$ is the proportion of the booster cohort *Z in the S* compartment out of the total proportion of the booster cohort *Z in the S and R* compartments on day *t-1* in age group *j* |

| |
|---|
| $p^{Z,R}_{t-1,j}$ is the proportion of the booster cohort *Z in the R* compartment out of the total proportion of the booster cohort *Z in the S and R* compartments on day *t-1* in age group *j*<br><br>Note: $p^{Z,S}_{t-1,j} + p^{Z,R}_{t-1,j} = 1$<br><br>$\omega^{Z}_{t,j}$ is the natural immunity waning rate on day *t* in age group *j* in cohort Z (*X*, *V*, *B, B2*, *B3*, or *B4)*<br><br>$\epsilon_{t,j}$ is the proportion of external cases on day *t* in age group *j* |

#### 5.2. Force of infection (unvaccinated)

$$\lambda^{*}_{t,i} = (Overall\ Scaling\ Factor_t) \times \left[\beta_t \sum_{j=1}^{9} \sum_{Z=\{X,V,B\}} c_{ij} I^{Z}_{t,j}\right]$$

$\lambda^{*}_{t,i}$ is the age-group specific force of infection at time *t* for age group *i*

$Overall\ \ Scaling\ Factor_t$ at time *t* is defined in section 1.2.2.

$\beta_t$ is the transmissibility parameter at time *t*

$c_{ij}$ is the rate at which individuals in age group *i* make contact with those in age group *j*

$I^{Z}_{t,j}$ represents the infectious individuals in cohort *Z* (*X*, *V*, *B, B2*, *B3*, or *B4*).at time *t* for age group *j*.

#### 5.3. Force of infection (vaccinated)

For the vaccinated cohorts, the force of infection calculation is adjusted based on the VE in the cohort:

$$\lambda^{Z}_{t,i} = \left(1 - VE^{X}_{t,i}\right) \times (Overall\ Scaling\ Factor_t) \times \left[\beta_t \sum_{j=1}^{9} \sum_{Z=\{X,V,B\}} c_{ij} I^{Z}_{t,j}\right]$$

$\lambda^{Z}_{t,i}$ is the age-group specific force of infection at time *t* for age group *i* in cohort *Z* (*X*, *V*, *B, B2*, *B3*, or *B4*).

$VE_{t,i}^{Z}$ is the vaccine effectiveness at time *t* for age group *i* in cohort *Z* (*X*, *V*, *B, B2*, *B3*, or *B4*).The vaccine effectiveness at time *t* is defined in the next section.

### 5.4. Daily vaccine effectiveness calculations

If no one is vaccinated in the cohort and age group, a VE value of zero is assumed (i.e., when day $t$ is less than the day of the beginning of the vaccination period). Once people have been vaccinated in the vaccination cohort and age group, the average vaccine effectiveness on day *t* is calculated as:

$$VE_{t,i}^{Z} = \frac{\left[\begin{array}{c}(Number\ newly\ vaccinated\ on\ day\ t \times initial\ VE) + \\ \left((VE\ Drop_t)\left(Number\ previously\ vaccinated\ on\ day\ t \times (VE_{t-1,i}^{Z} - daily\ waning\ rate\right)\right)\end{array}\right]}{[Total\ number\ in\ cohort\ Z\ on\ day\ t]}$$

$VE_{t,i}^{Z}$ is the vaccine effectiveness at time *t* for age group *i* in cohort *Z* (*X*, *V*, *B, B2*, *B3* or *B4*).

If the term $\left(VE_{t-1,i}^{Z} - daily\ waning\ rate\right)$ falls below zero, we assume a value of zero instead.

The term $(VE\ Drop_t)$ represents a drop in the vaccine effectiveness when a new strain with immune escape enters the population (see Section 1.3.2. There are only a few days in which this drop occurs over the course of the time horizon: if a new variant with immune escape emerges, the impact is assumed to happen on the individual days described in Section 5.5.8. Apart from those days, the value of the term $(VE\ Drop_t)$ is assumed to be one, corresponding to no impact on the average VE calculation.

### 5.5. Calculation of incremental effectiveness against hospitalization

For each vaccination cohort, age group, and day, we define the following vaccine effectiveness variables and relationship between the variables. The superscripts and subscripts for vaccination cohort, age group, and day are removed for clarity.

**Definitions**

$VE_1$ = Vaccine effectiveness against infection

$VE_2$ = 'Total' Vaccine effectiveness against hospitalization

$VE_2^{*}$= 'Additional' Vaccine effectiveness against hospitalization

We assume $VE_2^* = 0$ if there is no additional benefit against hospitalization

**Define**

$$[1 - VE_2] = [1 - VE_1] \times [1 - VE_2^*]$$

Isolate and solve for $VE_2^*$

$$[1 - VE_2^*] = \frac{[1 - VE_2]}{[1 - VE_1]}$$

$$VE_2^* = 1 - \frac{[1 - VE_2]}{[1 - VE_1]}$$

**Probabilities in an unvaccinated cohort**

Probability of COVID-19 infection in an unvaccinated cohort

$$p(COVID|UnVac)$$

Probability that a COVID-19 infection requires hospitalization in an unvaccinated cohort

$$p(Hosp|COVID, UnVac)$$

Proportion of an unvaccinated cohort with a COVID-19 infection that requires hospitalization

$$p(Hosp, COVID|UnVac) = p(Hosp|COVID, UnVac) \times p(COVID|UnVac)$$

**Probabilities in a vaccinated cohort**

Probability of COVID-19 infection in a vaccinated cohort

$$p(COVID|Vac) = [1 - VE_1] \times p(COVID|UnVac)$$

Probability that an COVD-19 infection requires hospitalization in a vaccinated cohort

$$p(Hosp|COVID, Vac) = [1 - VE_2^*] \times p(Hosp|COVID, UnVac)$$

Proportion of a vaccinated cohort with an COVID infection that requires hospitalization

$$p(Hosp, COVID|Vac) = p(Hosp|COVID, Vac) \times p(COVID|Vac)$$

### 6. REFERENCES

1. Kohli MA, Maschio M, Lee A, et al. The potential clinical impact of implementing different COVID-19 boosters in fall 2022 in the United States. *J Med Econ.* 2022;25(1):1127-1139.

2. United Nations Department of Economic and Social Affairs PD. World Population Prospects 2022, Online Edition. Population by Single Age – Both Sexes. https://population.un.org/wpp/Download/Standard/Population/. Published 2022. Accessed August 2, 2022.

3. Prem K, Cook AR, Jit M. Projecting social contact matrices in 152 countries using contact surveys and demographic data. *PLoS Comput Biol.* 2017;13(9):e1005697.

4. Institute for Health Metrics Evaluation (IHME). COVID-19 Projections. United States of America. Used with permission. All rights reserved. https://covid19.healthdata.org/united-states-of-america. Accessed May 31,, 2022.

5. Keeling MJ, Rohani P. *Modeling Infectious Diseases in Humans and Animals. Page 159.* Princeton, NJ: Princeton University Press; 2008.

6. Keeling MJ, Dyson L, Tildesley MJ, Hill EM, Moore S. Comparison of the 2021 COVID-19 roadmap projections against public health data in England. *Nature Communications.* 2022;13(1):4924.

7. Center for Disease Control and Prevention. COVID-19 Vaccination Demographics in the United States, National. https://data.cdc.gov/Vaccinations/COVID-19-Vaccination-Demographics-in-the-United-St/km4m-vcsb. Accessed: April 28, 2023.

8. Institute for Health Metrics and Evaluation (IHME). COVID-19 model update: Omicron and waning immunity. Available at: www.healthdata.org. Updated: December 22, 2021. Accessed: December 23, 2021.

9. Voko Z, Kiss Z, Surjan G, et al. Effectiveness and Waning of Protection With Different SARS-CoV-2 Primary and Booster Vaccines During the Delta Pandemic Wave in 2021 in Hungary (HUN-VE 3 Study). *Front Immunol.* 2022;13:919408.

10. Pratama NR, Wafa IA, Budi DS, et al. Effectiveness of COVID-19 Vaccines against SARS-CoV-2 Omicron Variant (B.1.1.529): A Systematic Review with Meta-Analysis and Meta-Regression. *Vaccines.* 2022;10(12):2180.

11. Higdon MM, Baidya A, Walter KK, et al. Duration of effectiveness of vaccination against COVID-19 caused by the omicron variant. *The Lancet Infectious diseases.* 2022;22(8):1114-1116.

12. Khoury DS, Cromer D, Reynaldi A, et al. Neutralizing antibody levels are highly predictive of immune protection from symptomatic SARS-CoV-2 infection. *Nature medicine.* 2021;27(7):1205-1211.

13. Hogan AB, Wu SL, Dooha P, et al. *Imperial College COVID-19 response team. Report 48: The value of vaccine booster doses to mitigate the global impact of the Omicron SARS-CoV-2 variant.* *https://www.imperial.ac.uk/mrc-global-infectious-disease-analysis/covid-19/report-48-global-omicron/.* 16 December 2021.

14. Chalkias S, Eder F, Khetan S, et al. Safety, immunogenicity and antibody persistence of a bivalent beta-containing booster vaccine. 15 April 2022, PREPRINT (Version 1) available at Research Square [https://doi.org/10.21203/rs.3.rs-1555201/v1]. 2022.

15. Chalkias S, Harper C, Vrbicky K, et al. A Bivalent Omicron-Containing Booster Vaccine against Covid-19. *N Engl J Med.* 2022;387(14):1279-1291.

16. Swanson K. *Pfizer/BioNTech COVID-19 Omicron-Modified Bivalent Vaccine. Presentation Slides. ACIP Meeting September 1, 2022.* Atlanta, GA, USA: Centers for Disease Control and Prevention.

17. Tseng H, Ackerson BK, Sy LS, et al.,. Effectiveness of mRNA-1273 bivalent (Original and Omicron BA.4/BA.5) COVID-19 vaccine in preventing hospitalizations for COVID-19, medically attended SARS-CoV-2 infections, and hospital death in the United States. *medRXiv.* 2023.

18. Kopel H, Nguyen VH, Boileau C, et al. Comparative Effectiveness of the Bivalent (Original/Omicron BA.4/BA.5) mRNA COVID-19 Vaccines mRNA-1273.222 and BNT162b2 Bivalent in Adults in the United States. *medRxiv.* 2023:2023.2007.2012.23292576.

19. Centers for Disease Control and Prevention, . COVID Data Tracker. https://covid.cdc.gov/covid-data-tracker/#vaccinations. 2023.

20. Wolfel R, Corman VM, Guggemos W, et al. Virological assessment of hospitalized patients with COVID-2019. *Nature.* 2020;581(7809):465-469.

21. Imai N, Cori A, Dorigatti I, et al. Report 3: Transmissibility of 2019-nCOV. Imperial College London. (25-01-2020). https://doi.org/10.25561/77148. Published 2020. Updated January 25, 2020. Accessed May 15, 2022.

22. Institute for Health Metrics and Evaluation (IHME). COVID-19 Projections. United States of America. https://covid19.healthdata.org/united-states-of-america. . Accessed January 23, 2022.

23. Vanni T, Karnon J, Madan J, et al. Calibrating models in economic evaluation: a seven-step approach. *Pharmacoeconomics.* 2011;29(1):35-49.

24. Covid-19 ForecastHub. Ensemble model. Available at: https://covid19forecasthub.org/doc/ensemble/. Accessed.

25. IHME. Covid-19 publications. Available at: https://www.healthdata.org/covid/publications. Accessed.

26. Centers for Disease Control and Prevention. COVID-19-Associated Hospitalization Rates (per 100,000 population) for the COVID-NET Network https://covid.cdc.gov/covid-data-tracker/#covidnet-hospitalization-network. Accessed May 19, 2023.

27. United States Bureau of Labor Statistics. Labor Force Statistics From the Current Population Survey. Employment status of the civilian noninstitutional population by age, sex, and race. https://www.bls.gov/cps/cpsaat03.pdf. Accessed November 20, 2022.

28. Centers for Disease Control and Prevention. National Center for Health Statistics. Long COVID. Household Pulse Survey. https://www.cdc.gov/nchs/covid19/pulse/long-covid.htm. Accessed November 20, 2022.

29. Ham DI. Long-Haulers and Labor Market Outcomes. Institute Working Paper 60. https://doi.org/10.21034/iwp.60. In:2022.

30. United States Bureau of Labor Statistics. Economic News Release: Employment Situation. Table B-3. Average hourly and weekly earnings of all employees on private nonfarm payrolls by industry sector, seasonally adjusted. https://www.bls.gov/news.release/empsit.t19.htm#. Accessed November 20, 2022.

31. IQVIA Institute Report. Trends in Vaccine Administration in the United States. https://www.iqvia.com/insights/the-iqvia-institute/reports/trends-in-vaccine-administration-in-the-united-states. Published 2023. Accessed August 7, 2023.

32. Prosser LA, Harpaz R, Rose AM, et al. A Cost-Effectiveness Analysis of Vaccination for Prevention of Herpes Zoster and Related Complications: Input for National Recommendations. *Ann Intern Med.* 2019;170(6):380-388.

33. Centers for Disease Control and Prevention. Isolation and Precautions for People with COVID-19. https://www.cdc.gov/coronavirus/2019-ncov/your-health/isolation.html. Accessed December 21, 2022.

34. Di Fusco M, Shea KM, Lin J, et al. Health outcomes and economic burden of hospitalized COVID-19 patients in the United States. *J Med Econ.* 2021;24(1):308-317.

35. Chopra V, Flanders SA, O'Malley M, Malani AN, Prescott HC. Sixty-Day Outcomes Among Patients Hospitalized With COVID-19. *Ann Intern Med.* 2021;174(4):576-578.

36. Moderna. Data on file. Summary of Solicited Adverse Reactions within 7 days after Booster injection by Grade solicity safety set (Part B, Open-Label Phase).

37. Klein N. Rapid cycle analysis to monitor the safety of COVID-19 vaccines in near real-time within the vaccine safety datalink: Myocarditis and anaphylaxis. Presentation to the Advisory Committee on Immunization Practices. August 30, 2021. https://www.cdc.gov/vaccines/acip/meetings/downloads/slides-2021-08-30/04-COVID-Klein-508.pdf. Accessed.

38. Shimabukuro T. Update on myocarditis following mRNA COVID-19 vaccination. Presentation to the Vaccines and Related Biological Products Advisory Committee. https://www.fda.gov/media/159007/download. Accessed June 7, 2022.

39. Center for Medicare and Medicaid Services. 2022 National Physician Fee Schedule Relative Value File October Release. https://www.cms.gov/medicare/medicare-fee-for-service-payment/physicianfeesched/pfs-relative-value-files. Published 2022. Accessed May, 2023.

40. Drugs.com. Acetaminophen/codeine Prices, Coupons and Patient Assistance Programs. https://www.drugs.com/price-guide/acetaminophen-codeine#:~:text=Acetaminophen%2F%20codeine%20Prices.%20The%20cost%20for%20acetaminophen%2Fcodeine%20oral,only%20and%20are%20not%20valid%20with%20insurance%20plans. Published 2021. Accessed October 18, 2021.

41. Centers for Disease Control and Prevention. Allergic Reactions Including Anaphylaxis After Receipt of the First Dose of Moderna COVID-19 Vaccine - United States, December 21, 2020 - January 10, 2021. *MMWR Morbidity and mortality weekly report.* 2021;70(4):125-129.

42. HCUPnet - Hospital Inpatient National Statistics. 2018 National Diagnoses - Clinical Classification Software Refined (CCSR), Principal Diagnosis: INJ031 Allergic Reactions. . https://hcupnet.ahrq.gov/#setup. Published 2018. Accessed October 13, 2021.

43. Centers for Medicare and Medicaid Services. Outpatient Prospective Payment System: Addendum B. - Final OPPS Payment by HCPCS Code for CY 2021. https://www.cms.gov/Medicare/Medicare-Fee-for-Service-Payment/HospitalOutpatientPPS/Addendum-A-and-Addendum-B-Updates. Published 2021. Accessed October 15, 2021.

44. Tutle KL WP. Capturing anaphylaxis through medical records. Are ICD and CPT codes sufficient. *Ann Allergy Asthma Immunol.* 2020;124:150-155.

45. HCUPnet - Hospital Inpatient National Statistics. 2018 National Diagnoses - Clinical Classification Software Refined (CCSR), Principal Diagnosis: CIR005 Myocarditis and Cardiomyopathy. . https://hcupnet.ahrq.gov/#setup. Published 2018. Accessed October 13, 2021.
